# The role of obesity in female reproductive conditions: A Mendelian Randomisation study

**DOI:** 10.1101/2021.06.01.21257781

**Authors:** Samvida S. Venkatesh, Teresa Ferreira, Stefania Benonisdottir, Nilufer Rahmioglu, Christian M. Becker, Ingrid Granne, Krina T. Zondervan, Michael V. Holmes, Cecilia M. Lindgren, Laura B. L. Wittemans

## Abstract

**Background:** Obesity is observationally associated with altered risk of many female reproductive conditions. These include polycystic ovary syndrome (PCOS), abnormal uterine bleeding, endometriosis, infertility, and pregnancy-related disorders. However, the roles and mechanisms of obesity in the aetiology of reproductive disorders remain unclear.

**Methods and Findings:** We estimated observational and genetically predicted causal associations between obesity, metabolic hormones, and female reproductive conditions using logistic regression, generalised additive models, and Mendelian randomisation (two-sample, non-linear, and multivariable) applied to data from UK Biobank and publicly available genome-wide association studies (GWAS).

Body mass index (BMI), waist-hip ratio (WHR), and WHR adjusted for BMI (WHRadjBMI) were observationally (odds ratios (ORs) = 1.02 – 1.87 per 1 S.D. obesity trait) and causally (ORs = 1.06 – 2.09) associated with uterine fibroids (UF), PCOS, heavy menstrual bleeding (HMB), and pre-eclampsia. Causal effect estimates of WHR and WHRadjBMI, but not BMI, were attenuated compared to their observational counterparts. Genetically predicted visceral adipose tissue mass was causal for the development of HMB, PCOS, and pre-eclampsia (ORs = 1.01 - 3.38). Increased waist circumference also posed a higher causal risk (ORs = 1.16 – 1.93) for the development of these disorders and UF than did increased hip circumference (ORs = 1.06 – 1.10). Leptin, fasting insulin, and insulin resistance each mediated between 20% -50% of the total causal effect of obesity on pre-eclampsia. Reproductive conditions clustered based on shared genetic components of their aetiological relationships with obesity.

**Conclusions:** In this first systematic, large-scale, genetics-based analysis of the aetiological relationships between obesity and female reproductive conditions, we found that common indices of overall and central obesity increased risk of reproductive disorders to heterogenous extents, mediated by metabolic hormones. Our results suggest exploring the mechanisms mediating the causal effects of overweight and obesity on gynaecological health to identify targets for disease prevention and treatment.

## Introduction

Obesity is commonly understood as the excess accumulation of body fat which leads to increased health risks. In women, body mass index (BMI) is associated with increased prevalence of gynecological conditions, including excessive and abnormal menstrual bleeding (1, 2), endometriosis and uterine fibroids (UF) (3, 4), polycystic ovary syndrome (PCOS) (5, 6), complications of pregnancy such as pre-eclampsia and eclampsia (7), miscarriage (8, 9), and infertility (10, 11). These are often non-linear and heterogeneous relationships. While the risk of anovulatory infertility and recurrent miscarriages are highest in obese women, underweight women also have increased risk of infertility (9, 12). The association of BMI with endometriosis varies by disease severity, as women with advanced-stage endometriosis have lower BMI than those with minimal disease, and the inverse BMI-endometriosis association is stronger in women with infertility (13, 14). Finally, although the severity of PCOS and menstrual disorders increases with overall obesity, women presenting with these conditions are more likely to store fat in the abdominal region, regardless of their BMI (2, 5).

Observational epidemiological studies are limited in assessing causality, due to confounding and reverse causation. The Mendelian randomisation (MR) framework is a genetics-based instrumental variable approach that relies on the random and fixed assignment of genetic variants at conception to estimate the causal effect size of genetically predicted exposures on an outcome. MR has previously indicated causal effects of genetically predicted BMI on the development of some subtypes of ovarian cancer (odds ratio (OR) = 1.29 per 5 units of BMI) (15), endometrial cancer (OR = 2.06 per 5 units of BMI) (16), and PCOS (OR = 4.89 per 1 S.D. higher BMI) (17). However, the aetiological role of obesity and body fat distribution on many other female reproductive diseases has not been reported. It is especially relevant to investigate the effects of fat distribution, as there are intricate metabolic and endocrine links between adipose tissue and female reproductive organs. Yet, causal investigations of such relationships are lacking.

Leptin, which is a hormone secreted by adipocytes, and elevated in individuals with obesity, is increased in women with endometriosis, UF, and adverse pregnancy outcomes, even when adjusted for BMI (7, 18-21). Obesity-induced insulin resistance additionally increases the risk and severity of PCOS and pre-eclampsia by dysregulating steroid hormone and metabolic pathways (5, 22, 23). The dysregulation of sex hormones, including oestrogen and testosterone, is likely to play a role in the obesity-driven development of female reproductive disorders due to its close associations with body fat (22, 24). Yet, the causal impact of these factors in mediating the relationships between obesity and gynaecological diseases has not been detailed.

Here, we apply logistic regression, generalised additive models, two-sample, non-linear, and multivariable MR to dissect the relationships of overall obesity and body fat distribution with a range of female reproductive disorders, and investigate the mediating role of metabolic factors including leptin and insulin.

## Methods

### Observational associations in UK Biobank

UK Biobank (UKBB) is a prospective UK-based cohort study with approximately 500,000 participants aged 40-69 at recruitment on whom a range of medical, environmental, and genetic information is collected (25). We included 257,193 individuals self-identifying as females of white ancestry in UKBB in our analyses. Baseline measurements of BMI (total body weight (kg) / standing height^2^ (m^2^)) and waist-to-hip ratio (WHR) (waist circumference (cm) / hip circumference (cm)), and WHR adjusted for BMI (WHRadjBMI) were used to estimate general and central obesity, respectively. Cases of reproductive conditions were identified based on ICD9 and ICD10 primary and secondary diagnoses from hospital inpatient data, self-reported illness codes, and primary care records (**Table A in S1 Table**). We fitted logistic regression models to estimate the associations of BMI, WHR, and WHRadjBMI with prevalence of endometriosis (7,703 cases, 249,490 controls), heavy menstrual bleeding (17,229 cases, 239,964 controls), infertility (2,194 cases, 254,999 controls), self-reported stillbirth, spontaneous miscarriage or termination (81,102 cases, 176,091 controls), PCOS (746 cases, 256,447 controls), pre-eclampsia (2,242 cases, 254,951 controls), and uterine fibroids (19,192 cases, 238,001 controls). Case definitions for pre-eclampsia included eclampsia cases to capture cases in which the former may have developed into the latter. For each disease, individuals not included in the case group were used as controls. BMI, WHR and WHRadjBMI were adjusted for age, age-squared, assessment centre, and smoking status. The residuals were rank-based inverse normally transformed. Multiple testing correction was applied using the false discovery rate (FDR) to evaluate statistical significance while minimising false negatives (26).

We also tested associations without adjustment for smoking status, as it has previously been suggested that higher BMI increases risk of smoking (27) and adjustment for both could therefore induce collider bias. Adjustment for menopause status was not performed as up to 42% of women with reproductive disorders in UKBB report being unsure of their menopause status as compared to 16% of women who do not have a recorded history or presence of a reproductive condition (**Table A in S1 Table**).

To evaluate the presence of non-linear observational associations between obesity and each reproductive trait, fractional polynomial regression following the closed test procedure was performed using the mfp v1.5.2 R package (28). This algorithm tests for the presence of an overall association, the likelihood of non-linearity, and selects the best-fitting fractional polynomial function. We also fitted generalised additive models (GAM) to the same data, allowing for smoothing of the obesity trait with splines, using the mgcv 1.8-31 R package (29). All models were adjusted for age, age-squared, assessment centre, and smoking status. Model fits were compared with Akaike’s Information Criterion (AIC) (30).

### Two-sample Mendelian Randomisation

Genetic instruments for BMI, WHR, and WHRadjBMI were selected based on the sentinel variants at genome-wide significant loci (*P* < 5E-9) reported in the largest publicly available European ancestry GWAS of Genetic Investigation of ANthropometric Traits (GIANT) and UKBB (max N individuals = 806,801) (31). Similarly, genetic instruments for predicted visceral adipose tissue (VAT) mass (N individuals = 325,153) (32), waist circumference (N individuals = 462,166) and hip circumference (N individuals = 462,117) (33), and waist-specific and hip-specific WHR (N individuals = 18,330) (34) were selected based on the largest publicly available GWAS.

Three instrument weighting strategies were considered where sex-stratified GWAS results were available: (i) SNPs from combined-sexes GWAS with combined-sexes weights (effect sizes), (ii) combined-sexes SNPs with female-specific weights, or (iii) female-specific SNPs with female-specific weights. The method of female-specific SNPs with female-specific weights produced the strongest instruments as evaluated by F-statistics and was thus chosen for analysis (**Table B in S1 Table**). Additionally, due to concerns of ascertainment bias in UKBB (35, 36), sensitivity analyses with combined-sexes instruments (combined-sexes SNPs with combined-sexes weights) were also performed.

Associations of the genetic instruments for obesity traits with female reproductive diseases were obtained by performing a fixed-effect inverse-variance weighted meta-analysis of publicly available GWAS summary statistics from two large biobank projects - FinnGen and UKBB (37). The meta-analysis was performed using METAL (38) by matching the relevant ICD codes (**Table C in S1 Table**) for the following traits: infertility (4,996 cases, 421,223 controls), pre-eclampsia (2,711 cases, 480,373 controls), and uterine fibroids (21,835 cases, 456,551 controls). For endometriosis, summary statistics were obtained by request from a recent European ancestry GWAS (39) and meta-analysed as above with publicly available FinnGen and UKBB summary statistics (12,210 cases, 450,183 controls). For heavy menstrual bleeding (HMB) (9,813 cases, 210,946 controls), sporadic miscarriage, i.e. 1-2 miscarriages (50,060 cases, 174,109 controls), and multiple consecutive miscarriage, i.e. >= 3 consecutive miscarriages (750 cases, 150,215 controls), publicly available summary statistics were obtained from recent European ancestry GWAS that include UKBB individuals (3, 40). For PCOS, estimates were based on a fixed-effect inverse variance-weighted meta-analysis of published GWAS summary statistics (38), publicly available GWAS results by FinnGen, and a European-ancestry GWAS run in UKBB using SAIGE (11,186 cases, 273,812 controls). As a sensitivity analysis, all MR tests were performed using disease association estimates based on FinnGen only, where available, to alleviate bias due to sample overlap between the exposure and outcome GWAS sources.

Power to detect MR associations was calculated using two methods, one designed for general two-sample MR (41) and the other for MR performed on binary outcomes (42). Briefly, these methods calculate power by accounting for GWAS sample size, proportion of cases in case-control GWAS, and variance explained by genetic instruments for the exposure. The power to detect a true odds ratio (OR) association of 1.1 or more extreme at an unadjusted significance level of 0.05 was estimated.

Instrument SNPs were extracted from the outcome GWAS results, harmonised for consistency in the alleles, and MR was performed using the TwoSample MR v0.5.4 R package (43). Three methods for MR -inverse-variance weighted (IVW), MR-Egger, and weighted median - were evaluated, and the best method was selected via Rucker’s framework (44). Briefly, this framework advises to choose the MR method with least heterogeneity as assessed by Cochran’s Q-statistic, while accounting for the trade-off between power and pleiotropy (45). Inverse-variance weighted results, which were the best method chosen by Rucker’s framework for all tested associations, are reported in the main text, but results from all methods are calculated for robustness and displayed in the supplementary information. Multiple-hypothesis testing correction was applied with the FDR method and significance established at FDR < 0.05. MR-Egger intercept tests were performed to detect horizontal pleiotropy, and single-SNP and leave-one-out analyses were used to identify outlier SNPs driving relationships (43).

Reverse MR for obesity traits regressed on female reproductive conditions was performed as detailed above. Genetic instruments for endometriosis (14,926 cases, 189,715 controls) (46), PCOS (10,174 cases, 103,164 controls) (47), and uterine fibroids (20,406 cases, 223,918 controls) (3) were constructed from index variants identified by the largest European ancestry GWAS for each trait. Instrument strength was assessed by F-statistics (endometriosis, 16 SNPs, F = 5.13; PCOS, 14 SNPs, F = 41.6; UF, 29 SNPs, F = 11.1). Associations of genetic instruments for these reproductive conditions with BMI, WHR, and WHRadjBMI were obtained from female-specific summary statistics from the above-mentioned GIANT-UKBB meta-analysis (31).

### Non-linear Mendelian Randomisation

For non-linear MR analyses, we selected female UK Biobank participants of white British ancestry with no second-degree or closer relatives in the study, as identified by the UKBB team (48), to avoid violation of the MR assumption of random assignment of genetic variants; 207,705 women were retained following this selection. Genetic instruments for BMI were constructed for each individual using female-specific index variants from Pulit et al.’s GIANT-UKBB meta-analysis (31). The instruments for BMI explained 4.15% of trait variance after adjustment for age, age-squared, smoking status, assessment centre, genotyping array, and the first ten genetic principal components to account for population stratification. Binomial non-linear MR, a method designed to assess causal relationships in different exposure strata while avoiding collider bias, was performed using the fractional polynomial method with 100 quantiles and the piecewise linear method with 10 quantiles (49). Outcomes were restricted to female reproductive disorders with prevalence > 5% in UKBB (i.e. HMB, miscarriage, and UF) to maintain sufficient sample sizes in each quantile to estimate localised average causal effects. We assessed non-linearity with the fractional polynomial non-linearity and Cochran’s Q tests, and tested heterogeneity of the instrumental variable (IV) with the Cochran’s Q and trend tests. All analyses were performed with the nlmr v2.0 R package (49).

### MR with Mediation Analysis

To investigate the extent to which obesity affects female reproductive disorders via hormone-related mediators, two-step MR by the product of coefficients method was performed using GWAS summary statistics. This method was chosen as female reproductive disease phenotypes are binary outcomes with disease prevalence < 10% in UK Biobank, for which two-step MR provides the least biased estimates of mediation (50). Summary statistics for leptin (N = 33,987) (51), fasting insulin (N = 51,750) (52), and insulin sensitivity (N = 16,753) (53) were obtained from publicly available European-ancestry GWAS sources that do not include samples from UKBB to minimise bias from sample overlap (**Table B in S1 Table**).

In the first step of two-step MR, the mediators were regressed on obesity-related exposures using summary statistics MR methods described above. The direction of causality for all relationships was confirmed with the MR-Steiger directionality test (54) and reciprocal MR with mediator instruments and obesity-related exposures as outcomes were performed to ensure correct direction of causality. In the second step, multivariable MR (MVMR) was performed using combined genetic instruments for each obesity trait and hormone to estimate the independent effect of the mediator on each outcome after adjusting for the value of the exposure; and to estimate the independent effect of the exposure on outcome when adjusted for the value of each mediator. This was only done for traits where the total unadjusted effect of the exposure on the outcome was significant (FDR < 0.05). Odds ratios (ORs) for binary outcomes were converted to log ORs to calculate mediated effect by the product of coefficients method. The proportion of effect mediated was calculated by dividing indirect effect over total effect. Standard errors were estimated with the delta method (55).

### Disease and SNP Clustering

To assess similarities in the aetiological relationships of different reproductive conditions with obesity traits, we projected single SNP causal estimates for BMI, WHR and WHRadjBMI on the reproductive traits, estimated using the Wald ratio, in a two-dimensional space using UMAP. SNPs were annotated to their nearest gene with SNPsnap (56).

To identify the genetic instruments driving the causal obesity-reproductive trait association, and identify clusters of SNPs with distinct causal effect sizes, we clustered SNPs by the magnitude of their causal effect using mixture model clustering in the MR-Clust v0.1.0 R package (56). For each obesity trait-reproductive disease pair, the algorithm distinguishes the genetic instruments for the obesity traits that do not have an effect on the disease (“null cluster”), from those which have a similar scaled effect on the disease (the “substantial clusters”), and those that have a scaled effect that cannot be grouped with other variants (“junk cluster”).

### Code availability

All scripts used in analyses are deposited at https://github.com/lindgrengroup/obesity_femrepr_MR

## Results

### Obesity traits are observationally associated with female reproductive diseases in UK Biobank

BMI at baseline assessment (age 40-69) was positively associated with the prevalence of most female reproductive disorders in UKBB, with the strongest association observed between BMI and pre-eclampsia (odds ratio (OR) per 1 S.D. higher BMI = 1.87, *P* = 1.90E-64). Associations with WHRadjBMI were null or lower than those for WHR (ORs for WHRadjBMI v. WHR for PCOS = 1.06 v. 1.48, pre-eclampsia = 1.02 v. 1.13, endometriosis = 1.02 v. 1.08, HMB = 1.06 v. 1.14, and UF = 1.02 v. 1.08), indicating that BMI may be driving many of the associations between WHR and female reproductive diseases (**Figures 1 & 2, Table 1**). Infertility was the only disorder for which BMI (OR = 0.894, *P* = 2.16E-07) and WHR (OR = 0.927, *P* = 4.08E-04) were inversely associated with disease.

**Table 1:**
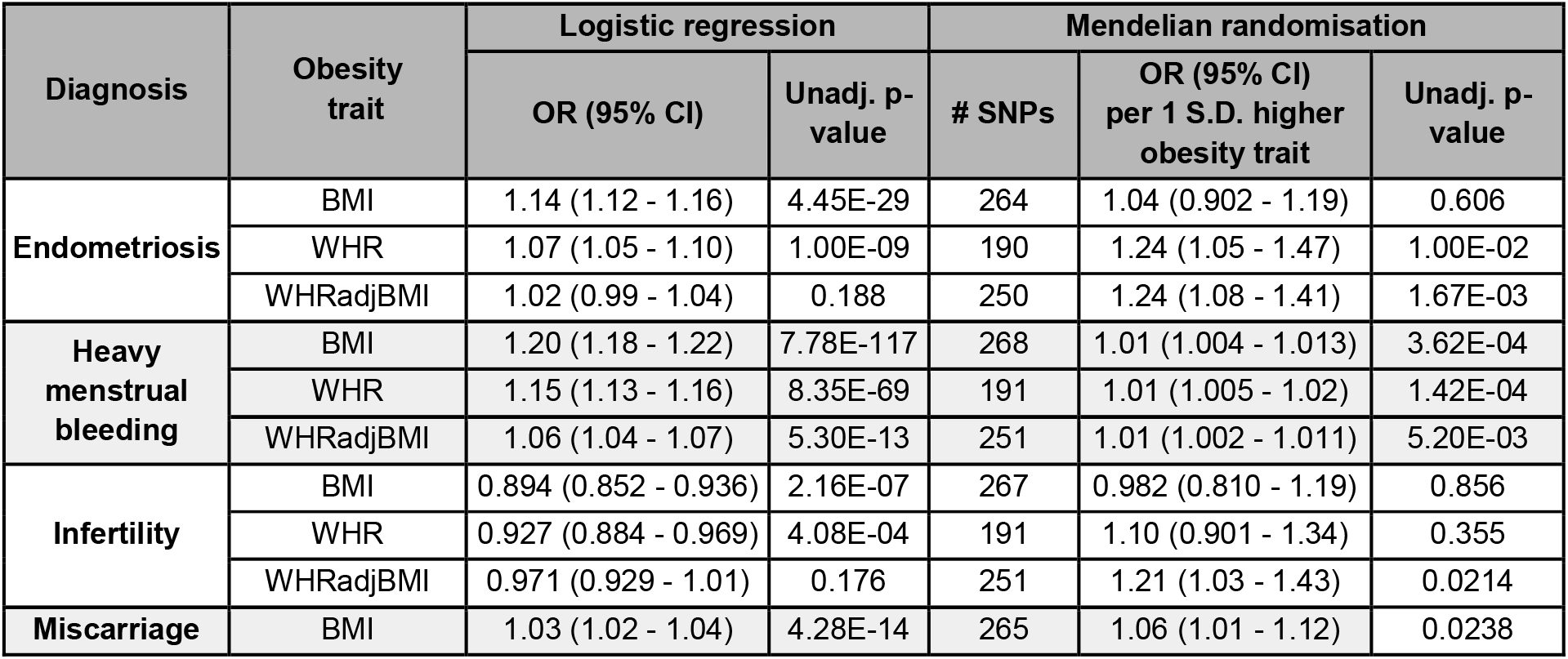
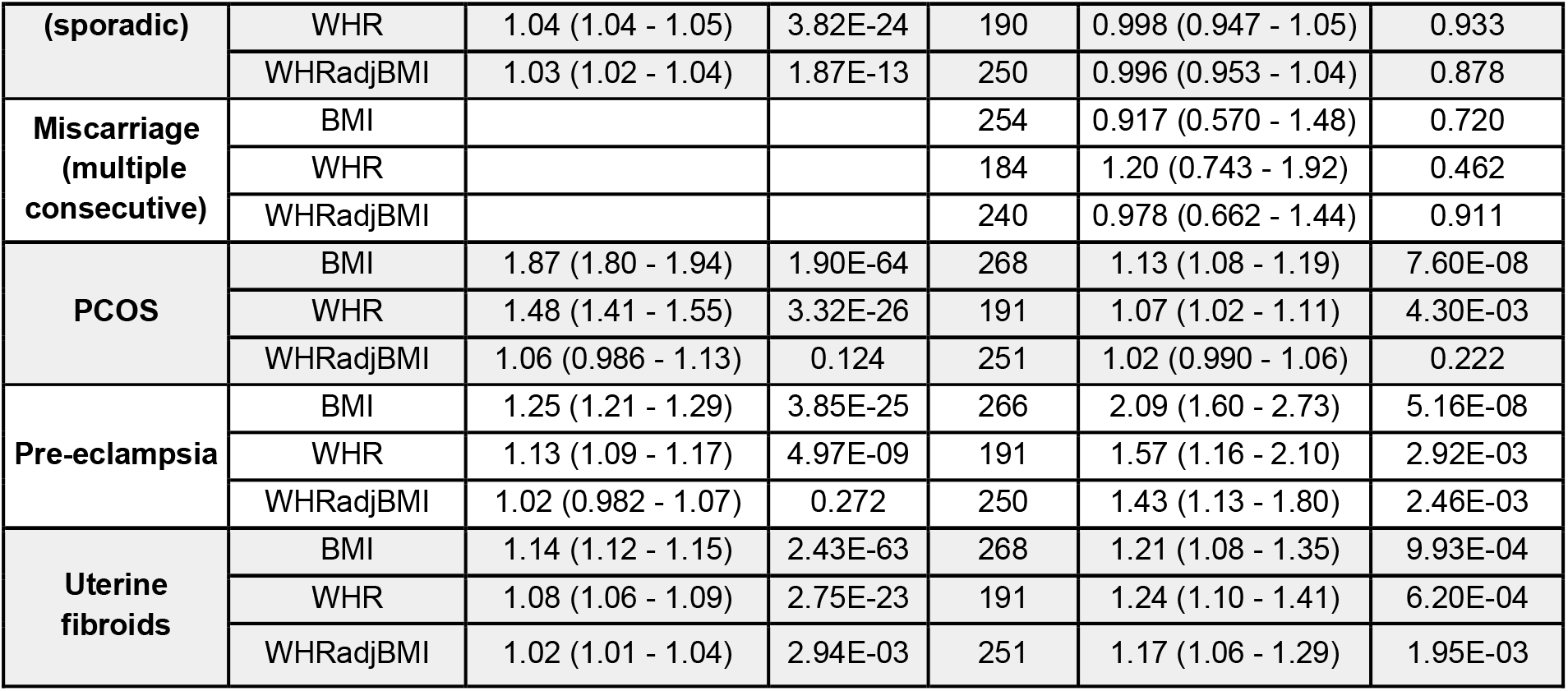
Observational and genetic associations between obesity traits and female reproductive disorders.

**Figure 1:**
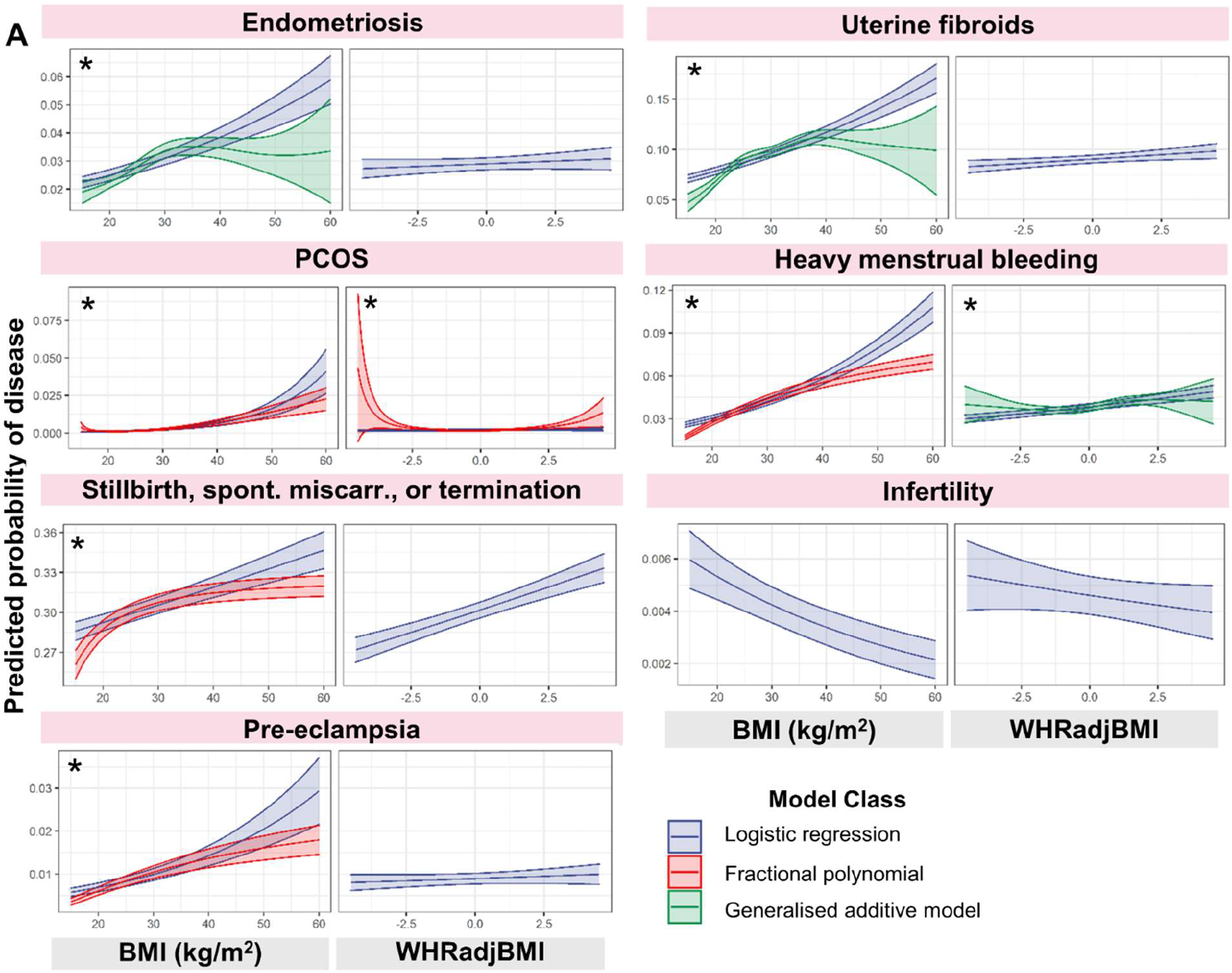
(A) Predicted probability of developing female reproductive disorders as a function of obesity-related traits. A series of logistic regression, fractional polynomial, and generalized additive models were fitted to estimate the probability of developing female reproductive disorders as a function of obesity-related traits in UKBB. Predicted fits for logistic and best-fitting non-linear models where better than the logistic regression (as evaluated with AIC) and 95% confidence intervals about the mean are displayed. * indicates that non-linear models fit the data better than linear models. Spont. miscarr. = spontaneous miscarriage.

**Figure 2:**
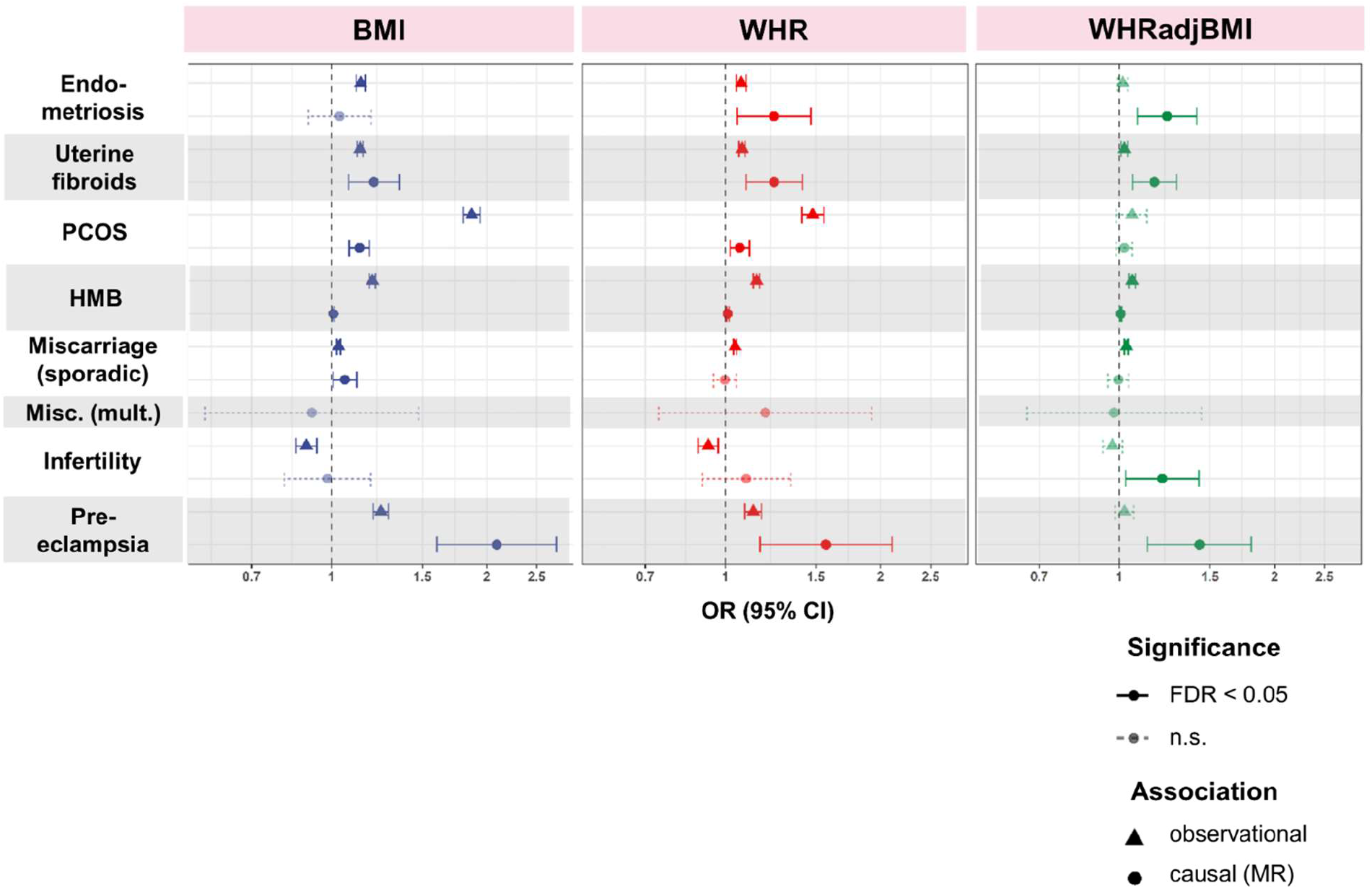
Comparison of observational and causal relationships between obesity-related traits and female reproductive disorders. Odds ratios (ORS) and 95% confidence intervals per 1 S.D. higher obesity trait displayed. Significant relationships (FDR adjusted p-value < 0.05) are in solid lines while non-significant (n.s.) ones are shown with dotted lines. For observational results, BMI, WHR, or WHRadjBMI adjusted for age, age-squared, region (assessment centre), and smoking status are used as predictors in a logistic regression model. Causal relationships between genetically predicted obesity-related traits and female reproductive disorders are assessed by two-sample Mendelian randomisation. The displayed method (inverse-variance weighted) is determined via Rucker’s model selection framework to minimise heterogeneity of the estimate. HMB = heavy menstrual bleeding, Misc. (mult.) = multiple consecutive miscarriage. Miscarriage (sporadic) = self-reported stillbirth, spontaneous miscarriage, or termination for observational results and sporadic miscarriage for MR results.

Non-linear models explained the associations of BMI with many reproductive disorders better than linear models. We observed inverted-U and plateau relationships with endometriosis (linear AIC = 67091, generalised additive model (GAM) AIC = 67051), uterine fibroids (linear AIC = 134160, GAM AIC = 134094), HMB (linear AIC = 116687, GAM AIC = 116636), miscarriage (linear AIC = 314828, GAM AIC = 314819), and pre-eclampsia (linear AIC = 24826, GAM AIC = 24814) (**Figure 1, Table D in S1 Table**). All three obesity traits displayed U-shaped relationships with PCOS.

Observational estimates between all obesity traits and female reproductive disorders did not differ with or without adjustment for smoking status (**Table E in S1 Table**). Statistical significance after multiple-testing correction was established at FDR < 0.05, unadjusted *P* < 0.04.

### Body fat distribution is causally related to risk of female reproductive diseases

Two-sample MR indicated that higher genetically predicted WHR and/or WHRadjBMI are causal for higher risk of pre-eclampsia (OR per 1 S.D. higher WHR = 1.57, *P* = 2.92E-03; WHRadjBMI = 1.43, *P* = 2.46E-03), endometriosis (WHR = 1.24, *P* = 1.00E-02; WHRadjBMI = 1.24, *P* = 1.67E-03), uterine fibroids (WHR = 1.24, *P* = 6.20E-04; WHRadjBMI = 1.17, *P* = 1.95E-03), infertility (WHRadjBMI = 1.21, *P* = 2.14E-02), and PCOS (WHR = 1.07, *P* = 4.30E-03) (**Table 1, Figure 2**). The causal estimates of WHR and WHRadjBMI on most reproductive disorders were higher than their observational counterparts. While genetically predicted BMI also increased risk of most female reproductive disorders (ORs per 1 S.D. higher BMI = 1.01 for HMB to 2.09 for pre-eclampsia), MR estimates of associations between BMI and HMB (OR = 1.01, *P* = 3.62E-04), endometriosis (OR = 1.04, *P* = 0.606), and PCOS (OR = 1.13, *P* = 7.60E-08) were much attenuated compared to observational results.

Genetically predicted visceral adipose tissue (VAT) mass was causal for the development of pre-eclampsia (OR per 1 kg increase in predicted VAT mass = 3.08, *P* = 6.65E-07), PCOS (OR = 1.15, *P* = 3.24E-05), and HMB (OR = 1.01, *P* = 0.0125) (**Figure 3** and **Table F in S1 Table**). The differential effect of body fat distribution on female reproductive traits was further reflected in the heterogeneous causal effects of waist circumference (WC) and hip circumference (HC) on disease development. Increased WC posed a higher risk than did increased HC for pre-eclampsia (ORs per 1 S.D. higher WC = 1.93 v. HC = 1.40, heterogeneity *P-het* = 0.0373), uterine fibroids (WC = 1.32 v. HC = 1.12, *P-het* = 7.70E-03), and PCOS (WC = 1.16 v. HC = 1.10, *P-het* = 0.0325). We did not see this heterogeneity in observational associations (all *P-het* > 0.164) (**Figure 3** and **Table F in S1 Table**).

**Figure 3:**
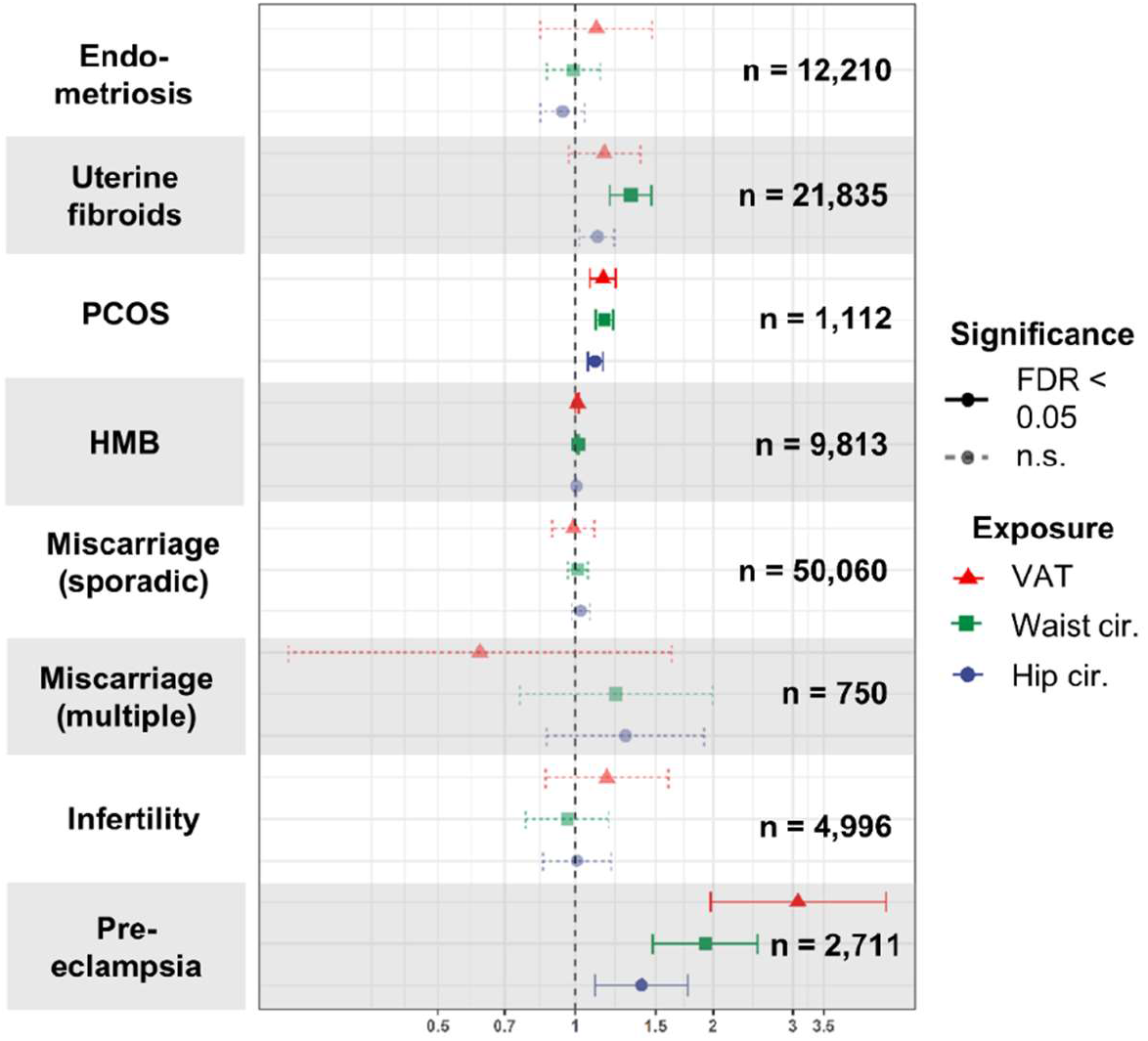
Causal relationships between genetically predicted visceral adipose tissue mass, waist circumference, hip circumference and female reproductive disorders. Odds ratios (ORs) per 1 kg increase in predicted visceral adipose tissue (VAT) mass or per 1 S.D. increase in waist circumference or hip circumference and 95% confidence intervals are estimated by two-sample MR. The displayed method (inverse-variance weighted) is determined via Rucker’s model selection framework to minimise heterogeneity of the estimate. Number of cases for each disease is indicated. Significant relationships (FDR adjusted p-value < 0.05) are in solid lines while non-significant ones are shown with dotted lines. Cir. = circumference, VAT = predicted visceral adipose tissue mass

No significant causal effects were found when restricting MR analyses to genetic instruments with a specific effect of waist but not hip circumference, or on hip but not waist circumference (**Table H in S1 Table** and **S1 Figure**), but the power based on these instruments to detect odds ratios more extreme than 1.1 was limited to 5% - 20% (**Table I in S1 Table**). No non-linear MR models explained the causal effects of BMI on any reproductive disorder better than linear MR models (**S3 Figure**). However, the power to detect non-linear effects was severely limited by the lower number of cases in each quantile of the BMI distribution in which analyses were run. Statistical significance after multiple-testing correction was established at FDR < 0.05, unadjusted *P* < 0.03.

SNPs identified in female-only GWAS and with female-specific weights for BMI, WHR, and WHRadjBMI (29) were found to be the strongest instruments, with F-statistics > 60; instrument strength for waist- and hip-circumference was > 45 (**Table B in S1 Table**). We found MR estimates to be consistent between the different MR methods (heterogeneity *P* > 0.321), when based only on FinnGen summary statistics (heterogeneity *P* > 0.163), or with combined-sex instruments (heterogeneity *P* > 0.999), suggesting that the findings were not dependent on the adopted MR method, or substantially biased due to sample overlap between exposure and outcome GWAS sources or ascertainment bias in UKBB (35, 36) (**Tables F, J, K in S1 Table & S1 & S2 Figure**).

We did not find evidence for reverse causal effects of endometriosis, PCOS, or uterine fibroids on BMI, WHR, and WHRadjBMI (**Table L in S1 Table**). However, these estimates may be biased by weak genetic instruments for endometriosis (F-statistic = 5.13) and UF (F-statistic = 11.1), and high heterogeneity for all associations (Cochran’s Q *P* < 4.71E-06). We were limited in assessing reverse causality of other female reproductive conditions on obesity traits by the lack of large-scale publicly available GWAS summary statistics.

### Leptin and insulin mediate the causal effects of obesity on female reproductive disorders

We applied a series of MR-based mediation analyses (58, 59) to study the role of hormonal factors - leptin and insulin resistance - in mediating the causal relationships between obesity and female reproductive health (**Figure 4A**). The effects of BMI, WHR, and WHRadjBMI on endometriosis, PCOS, pre-eclampsia, and UF were attenuated (95% CIs of ORs all contain 1) when adjusted for leptin, fasting insulin, or insulin sensitivity as measured by the modified Stumvoll Insulin Sensitivity Index (ISI) (**Figure 4B, Table M in S1 Table**). Furthermore, these hormones influence risk of pre-eclampsia independently of obesity. After adjustment for BMI, leptin (β = 0.887, *P* = 1.28E-04), fasting insulin (β = 1.42, *P* = 1.29E-03), and ISI (β = -0.503, *P* = 2.99E-03) were all associated with risk of pre-eclampsia (**Figure 4C, Table 2**). Similarly, fasting insulin (β = 1.27, *P* = 7.15E-03) and ISI (β = -0.793, *P* = 1.46E-05) had causal effects on pre-eclampsia upon adjustment for WHR. Leptin, fasting insulin, and ISI did not have significant causal effects on endometriosis, PCOS, or UF after adjustment for obesity traits. Statistical significance after multiple-testing correction was established at FDR < 0.05, unadjusted *P* < 0.01.

**Table 2:** Multivariable MR estimates of female reproductive disorders regressed on metabolic hormones, adjusted for obesity traits.

| Outcome | Exposure | Adjusted for | $\beta$ +/- S.E. per 1 S.D. higher exposure | Unadjusted P-value |
| --- | --- | --- | --- | --- |
| Endometriosis | Leptin | Unadjusted | -0.0745 +/- 0.188 | 0.692 |
|  |  | BMI | -0.0457 +/- 0.102 | 0.654 |
|  |  | WHR | -0.131 +/- 0.126 | 0.297 |
|  |  | WHRadjBMI | -0.0964 +/- 0.128 | 0.453 |
| Uterine fibroids | Leptin | Unadjusted | -0.132 +/- 0.323 | 0.683 |
|  |  | BMI | 0.0881 +/- 0.102 | 0.389 |
|  |  | WHR | -4.12E-03 +/- 0.138 | 0.976 |
|  |  | WHRadjBMI | -0.0834 +/- 0.14 | 0.550 |
| PCOS | Leptin | Unadjusted | -0.134 +/- 0.0701 | 0.0567 |
|  |  | BMI | 0.0491 +/- 0.0409 | 0.231 |
|  |  | WHR | 8.44E-03 +/- 0.0476 | 0.859 |
|  |  | WHRadjBMI | -0.0310 +/- 0.0473 | 0.511 |
| Pre-eclampsia | Leptin | Unadjusted | 0.181 +/- 0.501 | 0.718 |
|  |  | BMI | 0.887 +/- 0.232 | 1.28E-04 |
|  |  | WHR | 0.468 +/- 0.309 | 0.129 |
|  |  | WHRadjBMI | -0.102 +/- 0.333 | 0.760 |
| Endometriosis | Fasting insulin | Unadjusted | 0.150 +/- 0.247 | 0.544 |
|  |  | BMI | 0.0996 +/- 0.209 | 0.634 |
|  |  | WHR | 0.186 +/- 0.254 | 0.465 |
|  |  | WHRadjBMI | 0.423 +/- 0.239 | 0.0770 |
| Uterine fibroids | Fasting insulin | Unadjusted | 0.0263 +/- 0.342 | 0.939 |
|  |  | BMI | 0.311 +/- 0.183 | 0.090 |
|  |  | WHR | 0.313 +/- 0.209 | 0.135 |
|  |  | WHRadjBMI | 0.262 +/- 0.202 | 0.195 |
| PCOS | Fasting insulin | Unadjusted | -0.0334 +/- 0.158 | 0.833 |
|  |  | BMI | 0.0283 +/- 0.0772 | 0.714 |
|  |  | WHR | 0.0932 +/- 0.0788 | 0.237 |
|  |  | WHRadjBMI | 0.0289 +/- 0.0726 | 0.690 |
| Pre-eclampsia | Fasting insulin | Unadjusted | -0.0379 +/- 0.873 | 0.965 |
|  |  | BMI | 1.42 +/- 0.441 | 1.29E-03 |
|  |  | WHR | 1.27 +/- 0.473 | 7.15E-03 |
|  |  | WHRadjBMI | 0.476 +/- 0.467 | 0.308 |
| Endometriosis | Insulin sensitivity | Unadjusted | -0.0940 +/- 0.133 | 0.478 |
|  |  | BMI | -0.101 +/- 0.0864 | 0.244 |
|  |  | WHR | -0.131 +/- 0.100 | 0.192 |
|  |  | WHRadjBMI | -0.102 +/- 0.106 | 0.336 |
| Uterine fibroids | Insulin sensitivity | Unadjusted | -0.223 +/- 0.137 | 0.103 |
|  |  | BMI | -0.0951 +/- 0.0716 | 0.184 |
|  |  | WHR | -0.202 +/- 0.0830 | 0.0151 |
|  |  | WHRadjBMI | -0.120 +/- 0.0855 | 0.165 |
| PCOS | Insulin sensitivity | Unadjusted | -0.0239 +/- 0.0616 | 0.698 |
|  |  | BMI | -0.0257 +/- 0.0290 | 0.376 |
|  |  | WHR | -0.0355 +/- 0.0293 | 0.226 |
|  |  | WHRadjBMI | -0.0443 +/- 0.0294 | 0.132 |
| Pre-eclampsia | Insulin sensitivity | Unadjusted | -0.354 +/- 0.295 | 0.229 |
|  |  | BMI | -0.503 +/- 0.169 | 2.99E-03 |
|  |  | WHR | -0.793 +/- 0.183 | 1.461E-05 |
|  |  | WHRadjBMI | -0.519 +/- 0.205 | 0.0110 |

**Figure 4.**
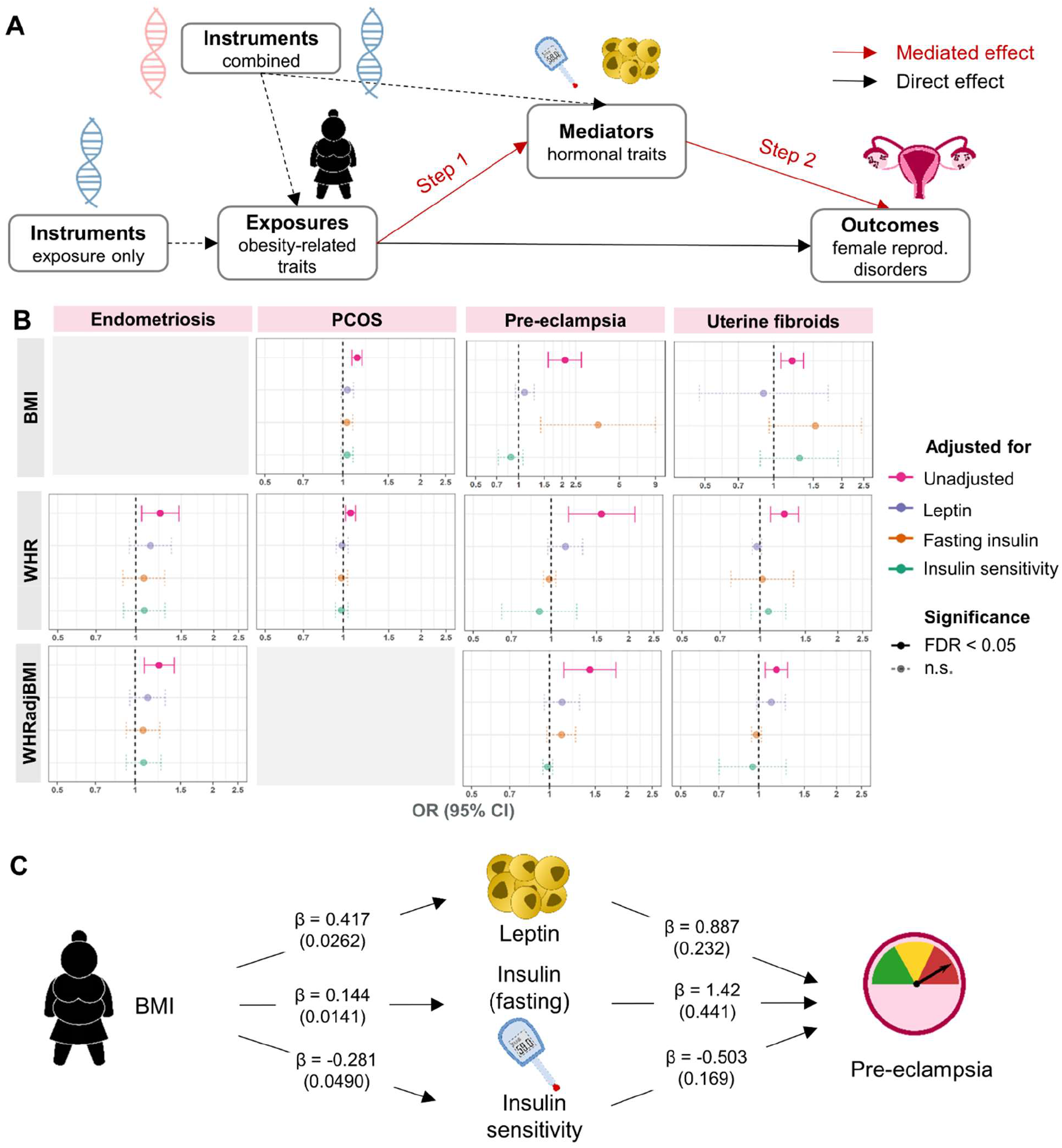
**(A) Method outline for two-step MR to estimate hormonally mediated effects between obesity and female reproductive disorders**. In step 1, the effect of exposures on mediators is estimated using instruments for the exposure alone, while in step 2 the independent effect of the mediators on outcomes is estimated using multivariable MR (MVMR) adjusted for exposures. All SNP-phenotype effect estimates come from different GWAS sources. **(B) MVMR estimates for obesity traits on female reproductive disorders adjusted for mediators**. MVMR was performed with combined genetic instruments for each exposure-mediator combination, displayed here for relationships where unadjusted exposure-outcome effect was significant (FDR < 0.05). **(C) Example of a mediated relationship**. Shown here for BMI effect on pre-eclampsia. Effect size estimates for exposure-mediator (betas and standard errors) and for mediator-outcome (log odds ratios and standard errors) are shown.

We calculated the proportion of total obesity effect mediated by the above hormones for disorders where the effects of obesity traits and mediators were significant at unadjusted *P* < 0.05. We found that leptin (50.2% of effect of BMI on pre-eclampsia), fasting insulin (between 27.7% - 36.6% of different effects), and ISI (between 19.1% - 50.1% of different effects) each mediated the total causal effect of obesity traits on female reproductive disorders (**Table 3**).

**Table 3:** Proportion of effect mediated for exposure - mediator - outcome relationships.

| Exposure | Mediator | Outcome | Log OR<br>(S.E.) per 1 S.D. higher exposure<br>*Unadj. pval* |  |  | Proportion of<br>Effect Mediated<br>(95% CI) |
| --- | --- | --- | --- | --- | --- | --- |
|  |  |  | Exposure -<br>Outcome | Mediator -<br>Outcome | Exposure -<br>Mediator |  |
| BMI | Leptin | Pre-eclampsia | 0.737<br>(0.135)<br>*P* = 5.16E-08 | 0.887<br>(0.232)<br>*P* = 1.28E-04 | 0.417<br>(0.0262)<br>*P* = 7.94E-57 | 50.2%<br>(18.2% - 82.2%) |
|  | Fasting insulin |  |  | 1.42<br>(0.441)<br>*P* = 1.29E-03 | 0.144<br>(0.0141)<br>*P* = 1.61E-24 | 27.7%<br>(7.40% - 48.0%) |
|  | Insulin sensitivity |  |  | -0.503<br>(0.169)<br>*P* = 2.99E-03 | -0.281<br>(0.0490)<br>*P* = 9.83E-09 | 19.1%<br>(3.33% - 35.0%) |
| WHR | Fasting insulin | Pre-eclampsia | 0.449<br>(0.151)<br>*P* = 2.92E-03 | 1.27<br>(0.473)<br>*P* = 7.15E-03 | 0.129<br>(0.0159)<br>*P* = 5.04E-16 | 36.6%<br>(0% - 73.7%) |
|  | Insulin sensitivity |  |  | -0.793<br>(0.183)<br>*P* = 1.46E-05 | -0.283<br>(0.0601)<br>*P* = 2.44E-06 | 50.1%<br>(4.98% - 95.3%) |
|  | Insulin sensitivity | Uterine fibroids | 0.218<br>(0.0637)<br>*P* = 6.20E-04 | -0.202<br>(0.083)<br>*P* = 1.51E-02 | -0.283<br>(0.0601)<br>*P* = 2.44E-06 | 26.2%<br>(0% - 54.4%) |
| WHRadjBMI | Insulin sensitivity | Pre-eclampsia | 0.358<br>(0.118)<br>*P* = 2.46E-03 | -0.519<br>(0.205)<br>*P* = 1.14E-02 | -0.168<br>(0.0427)<br>*P* = 8.21E-05 | 24.4%<br>(0% - 51.8%) |

### Other metabolic and hormone pathways may drive the aetiological relationships of obesity with female reproductive diseases

We assessed the similarities in the aetiological relationships of different reproductive conditions with obesity, by projecting the single SNP causal estimates for BMI, WHR and WHRadjBMI on the reproductive traits in a two-dimensional space using UMAP (**Figure 5A**). The UMAP projections based on all obesity traits clustered endometriosis and UF together with infertility and HMB, which were further separated from miscarriage (sporadic and multiple consecutive). While PCOS and pre-eclampsia were grouped closely in UMAP plots of the effect of WHR and WHRadjBMI variants, they were separated by BMI-associated variants. This reflects a shared genetic component of the aetiological role of general and central obesity in the three groups of reproductive conditions.

**Figure 5:**
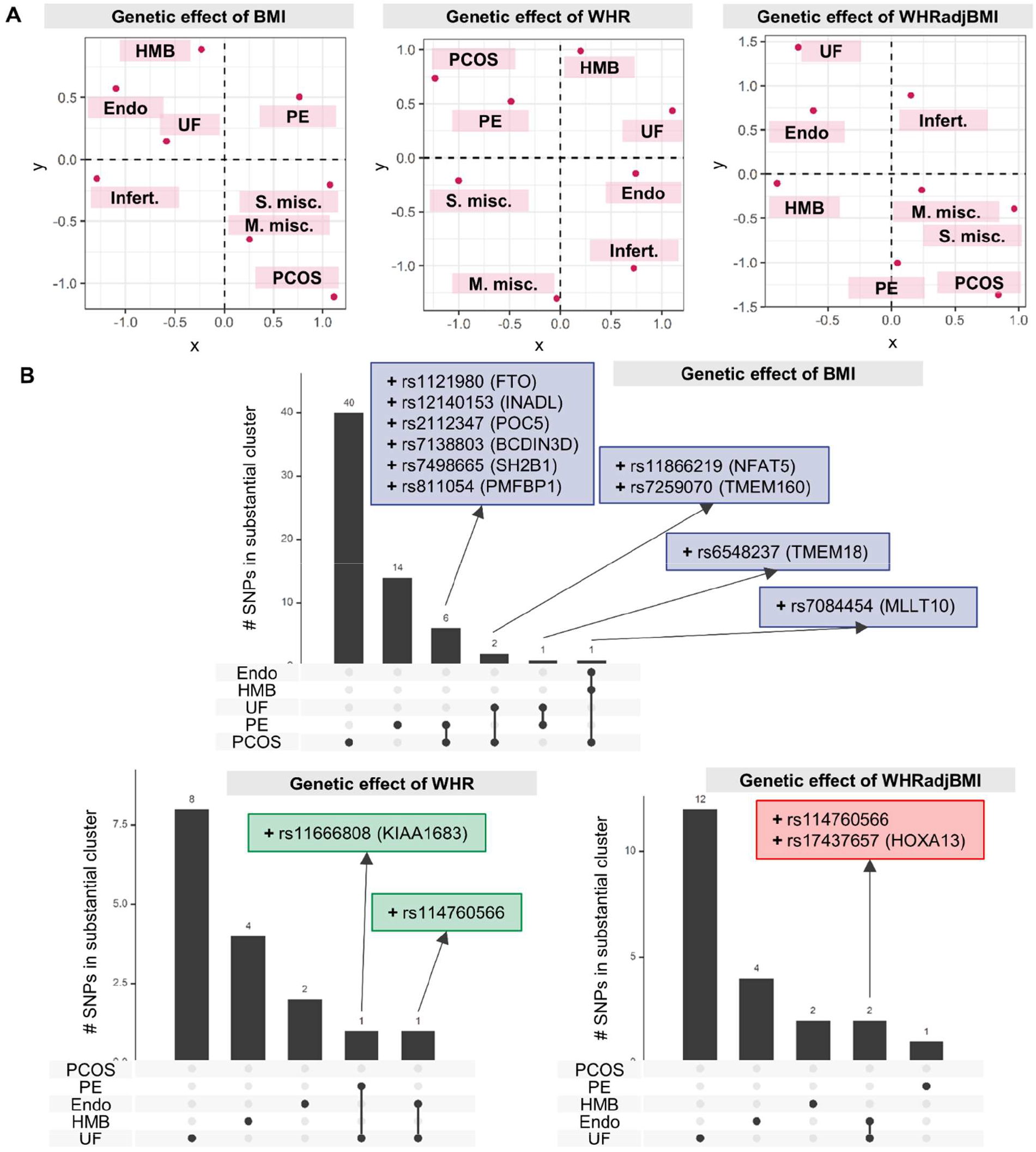
**(A) Female reproductive disorders separated by the genetic effects of different obesity traits on reproductive conditions**. Uniform Manifold Approximation and Projection (UMAP) was used to plot the separation of diseases based on single-SNP genetic effects of obesity instruments, evaluated by two-sample MR. **(B) Number of SNPs in substantial clusters for each obesity x female reproductive condition relationships**. SNPs with >= 80% probability of belonging to a substantial cluster (MRClust) are displayed with their nearest gene as annotated by SNPSnap and “+” or “-” representing causal effect direction. Infert = infertility, endo = endometriosis, UF = uterine fibroids, PE = pre-eclampsia, S. misc. = sporadic miscarriage, M. misc. = multiple consecutive miscarriage.

We further examined if different aspects of obesity play an aetiological role in different reproductive conditions. For each obesity trait-reproductive disease pair, we grouped the genetic instruments for the obesity traits by those that do not have an effect on the disease (“null cluster”), those which have a similar scaled effect on the disease (the “substantial clusters”), and those that have a scaled effect that cannot be grouped with other variants (“junk cluster”) using MRClust (57). One substantial cluster was identified for each pair of obesity traits and reproductive conditions. The only exception to this was with WHRadjBMI and UF, for which two substantial clusters were identified, one with positive causal effect and the other with negative effect (**S4 Figure** and **Table N in S1 Table**). Of the 4 SNPs in the negative effect cluster, rs2277339 (missense variant in *PRIM1* and upstream of *HSD17B6*, involved in steroid biosynthesis) is associated with primary ovarian insufficiency, early menopause, and PCOS (60, 61), and rs11694173 is intronic to *THADA*, which is also associated with PCOS (47). On the other hand, 4 of 10 SNPs in the positive effect cluster are associated with metabolic traits - rs12328675 and rs2459732 with circulating leptin (62), rs6905288 with type 2 diabetes and thyroid stimulating hormone (63, 64), and rs4686696 is intronic to insulin-like growth factor *IGF2BP2*.

SNPs with high probability of belonging to the substantial cluster (≥ 80% probability) were generally unique to each obesity-disease relationship, with no more than 2 variants shared between any two clusters (**Figure 5B**). However, 6 BMI index SNPs had positive causal effect estimates for both PCOS and pre-eclampsia, including rs1121980 in the adipose-associated gene *FTO* and rs7498665 in *SH2B1*, linked to insulin resistance in obesity. The BMI-associated variant rs7084454 (intronic to *MLLT10*) was shared by substantial clusters for PCOS, endometriosis, and UF, while rs114760566 (mapped to *HMGA1*, associated with type 2 diabetes and multiple lipomatosis) was shared by endometriosis and UF. We evaluated the biological effect of the top SNPs in each substantial cluster with the DEPICT algorithms for pathway enrichment and gene prioritisation. We recapitulated the known associations of GEMIN5 subnetwork enrichment in SNPs causal for BMI-PCOS, which has previously been implicated in the aetiology of PCOS (65). Gene prioritisation for WHR-endometriosis causal SNPs highlighted *TBX15*, an important mesodermal transcription factor with roles in endometrial and ovarian cancer (66, 67).

## Discussion

In this first systematic genetics-based causal investigation of the aetiological role of obesity in female reproductive health, we report evidence that common indices of obesity increase risk of a broad range of reproductive conditions whose effects may be non-uniform across the obesity spectrum. The strongest effect of generalised obesity was found for pre-eclampsia, while more modest effects were observed for nearly all other studied conditions. We identified endocrine mechanisms, including those related to leptin and insulin resistance, as potential drivers of aetiological relationships of both generalised and central obesity with female reproductive health. Finally, we found genetic evidence that certain groups of reproductive conditions, such as UF and endometriosis, may share a mechanistically similar relationship with obesity.

Our findings highlight that the relationships between obesity and female reproductive disorders are (i) non-uniform in their nature and strength, and (ii) observationally non-linear across the obesity spectrum. We report substantial differences in the causal effect estimates of BMI on reproductive diseases, with each S.D. increase in BMI doubling the risk of pre-eclampsia, but more moderately (ORs = 1.01 - 1.25 for PCOS, miscarriage, UF, HMB) or not at all (infertility, endometriosis) affecting other conditions. Conversely, central fat distribution independent of BMI showed substantial genetically predicted effects on both infertility and endometriosis (ORs per 1 S.D. increase in WHRadjBMI = 1.21 - 1.46) as well as on pre-eclampsia and UF (ORs = 1.17 - 1.43), but not on PCOS, HMB, and miscarriage. These findings highlight that the aetiological role of obesity in female reproductive diseases is heterogeneous in its effect strength, and may be driven by overall adiposity (PCOS, HMB, and miscarriage), isolated central obesity (infertility and endometriosis), or by both generalised and central obesity (pre-eclampsia and UF).

For several reproductive conditions, we found substantial differences between the observational and genetically predicted causal effect estimates, which may indicate a bi-directional relationship between obesity and reproductive health. For instance, while the observational analyses suggested an 87% increase in PCOS risk per S.D. higher BMI, the MR analyses indicated that each S.D. higher BMI causally increases PCOS risk by only 13%. Similarly, 1 S.D. higher WHR and WHRadjBMI causally increase endometriosis risk by 24%, while the observational analyses suggest a more modest increase in risk of 7% and 2% respectively. This discrepancy may in part be due to reverse causality, which we were not powered to detect in this study, as the number and strength of the available genetic instruments for reproductive conditions is substantially lower than those for BMI and WHR. The obesity traits upon which the observational analyses were based were measured at ages 44-67, which was for most conditions likely to be several years or decades after women developed the condition, and often post-menopause. While our observational analyses adjusted for the effect of age on obesity traits, adjusting for menopause status proved to be unreliable as up to 42% of women with reproductive diseases in UKB were unsure of their menopause status, as opposed to 16% of female participants without a diagnosis for any of the studied conditions. The observational estimates may therefore capture both the effect of obesity on disease risk as well as any downstream effects of the disease or commonly used treatments on body weight and fat distribution. For instance, the large observational effect of BMI on PCOS prevalence may reflect both a causal effect of obesity on disease risk (68), as captured by the genetically predicted effect, as well as weight gain as a consequence of PCOS (69). Other potential contributing factors to the differences between genetic and observational estimates are confounding by unmeasured variables which lead to inflated observational associations (70, 71), referral bias wherein obesity status affects the likelihood of receiving a diagnosis (72, 73), or differences in pre- and post-menopausal weight and body fat distribution not captured by age (74). Finally, while the observational relationships between obesity and some female reproductive disorders were non-linear, we did not find non-linearity in the causal effects of BMI on these diseases. The non-linear MR analyses were likely under-powered to detect associations with few cases in each quantile of the BMI spectrum.

We noted that genetic estimates for the effect of fat distribution were not similarly attenuated when compared to BMI effects. This disparity may be due to the differing impacts of overall and abdominal (central) adiposity, as the latter is thought to be biologically more directly linked to female reproductive health than generalised obesity, via pathways including insulin resistance and hyper-androgenaemia (5, 75-77). Supporting the stronger effect of central body fat, we also reported higher causal effects of waist than hip circumference with HMB, PCOS, pre-eclampsia, and UF. Genetically predicted visceral adipose tissue mass increased risk of PCOS and pre-eclampsia, in line with observational studies (78). VAT mass is also observationally associated with uterine fibroids (76), yet we did not find a significant causal effect of genetically predicted VAT mass on development of UF, which may suggest a bi-directional or reverse causal relationship.

Endometriosis and infertility were the only reproductive conditions which did not show a consistently positive link with obesity. The modest observational associations of both BMI and WHR with higher endometriosis prevalence in UKBB contradict previous studies, including prospective cohort studies, which reported that lower BMI was associated with increased disease prevalence (14, 79, 80). The positive association with endometriosis may in part be due to weight gain as a consequence of the disease, for instance due to hormonal treatments (81-83), chronic pain (84), inflammation (85), or earlier onset of menopause (86). We however did not find evidence that generalised obesity plays a causal role in the aetiology of endometriosis, which suggests that the observational finding reflects a reverse causal relationship. Conversely, the positive genetically predicted effect of WHRadjBMI on endometriosis risk indicates a causal role for abdominal fat distribution. For infertility, we observe a similar divergence between the observational and genetically predicted effects of obesity traits, with BMI showing a negative observational association, but WHRadjBMI a genetically predicted positive association. The causes of female infertility are multiple, ranging from PCOS (87) and anovulation (88), to tubal disease (89), endometriosis (90), low oocyte quality (91), hormonal and immunological dysfunction (92-95), and yet unknown mechanisms. Each of these may have distinct and complex relationships with obesity, which cannot be captured by studying the links with infertility of any cause. Nonlinear effects, such as the increased association of under-and overweight with incidence of infertility (12, 96), may also obscure these estimates, although our observational analyses did not provide evidence for a non-linear relationship.

We conducted the first genetics-based investigation of the mediating hormonal pathways underlying the causal relationships between obesity and female reproductive health. We identify mechanisms related to insulin resistance and leptin as mediators of the effects of obesity traits on UF and pre-eclampsia. The latter is consistent with hypotheses that obese women with metabolic dysregulation are at highest risk of developing hypertensive disorders of pregnancy via angiogenic and pro-inflammatory mechanisms. Increased circulating leptin may have a vasoconstructive, hypertensive effect, which may be worsened by attenuation of insulin-induced vasorelaxation and increased levels of TNF-alpha and IL6 (7, 21).

Finally, genetic clustering of female reproductive conditions revealed common genetic causes of obesity on endometriosis, UF, and HMB, which are known to share mechanisms of development (3, 97). The projection of infertility with these diseases merits following up on the genetic basis of endometriosis-related infertility with an eye to prevention and treatment.

The main strength of our work is the systematic approach to characterising the relationship between a broad range of obesity traits and common female reproductive conditions using both observational and genetic approaches. All observational associations were estimated in the same large-scale cohort study, which tends to lead to less biased estimates than case-control studies upon which most previous results were based. Moreover, we conducted the first genetics-based mediation analyses to pinpoint the mechanisms driving the causal effect of obesity on risk of reproductive diseases.

Reproductive conditions remain underdiagnosed and underreported in the UK, which was reflected in their low prevalence among female UKBB participants (**Table A in S1 Table**). This posed a limitation to our analyses in UK Biobank by reducing power to identify significant associations. For this reason, we opted to use broad case categories, such as infertility of any cause, as we had insufficient power and information to examine conditions by sub-types. Secondly, we restricted our analyses to women of genetically European ancestry, due to a lack of genetic data on women of other ancestries. Many of the reproductive diseases included here, with uterine fibroids being the most notable example (98, 99), are more prevalent in non-European populations and our results may not be transferable to women of other ancestries (100-102), which emphasises the urgent need to set up large-scale studies similar to UK Biobank on participants of non-European ancestry. We were further limited in investigations of metabolic, hormonal, and inflammatory mediating mechanisms by a lack of publicly available GWAS summary statistics for these traits. Finally, the lack of data on BMI and WHR prior to disease onset, and limited information on the age at which reproductive conditions were first diagnosed, complicated the interpretation of our findings from observational analyses in UK Biobank.

Key priorities for the future are the further exploration and validation of the pathways through which obesity increases risk of female reproductive disease. Notably, our finding that insulin resistance may be an important mediating mechanism warrants further attention, as cheap and safe treatments are available to increase insulin sensitivity. This is demonstrated by the successful use of metformin as a first-line treatment in women with PCOS (103), but such a treatment strategy has not yet been explored for other reproductive conditions linked to obesity. More generally, better and more detailed diagnostic information on reproductive health in large-scale cohort studies is urgently required for future research on the causes, consequences and aetiological mechanisms of female reproductive illnesses.

In conclusion, we provide genetic evidence that both generalised and central obesity play an aetiological role in a broad range of female reproductive conditions, but the extent of this link differs substantially between conditions. Our findings also highlight the importance of hormonal pathways, notably leptin and insulin resistance, as mediating mechanisms and potential targets for intervention in the treatment and prevention of common female reproductive conditions.

## Data Availability

All data referred to in the manuscript are publicly available summary statistics. All scripts used in analyses are deposited at: https://github.com/lindgrengroup/obesity_femrepr_MR

https://github.com/lindgrengroup/obesity_femrepr_MR

## Acknowledgements

We acknowledge the participants and investigators of FinnGen for their contribution to this study. This research has been conducted using the UK Biobank Resource under Application Number 10844. The views expressed are those of the author(s) and not necessarily those of the NHS, the NIHR or the Department of Health.

## Supporting Information Captions

**S1 Figure. Comparison of different Mendelian randomisation methods for regression of female reproductive diseases on various obesity traits**. Summary statistics MR was performed with the R package TwoSampleMR using three different methods whose results are compared. Effect sizes are displayed as odds ratios (ORs) with 95% confidence intervals (CIs). P-values are adjusted for multiple testing with the false discovery rate (FDR) correction; solid lines indicate associations that are significant at an adjusted p-value threshold of 0.05. BMI = body mass index, IVW = Inverse-variance weighted, PCOS = polycystic ovary syndrome, WHR = waist-hip ratio, WHRadjBMI = WHR adjusted for BMI, VAT = genetically predicted visceral adipose tissue mass.

**S2 Figure. Mendelian randomisation sensitivity analyses for female reproductive disorders regressed on obesity traits. (A)** Female-specific genetic instruments vs combined-sexes genetic instruments. **(B)** Reproductive outcomes from meta-analysis of UKBB and FinnGen summary statistics vs those from FinnGen only. Summary statistics MR performed with TwoSampleMR R package and best method (displayed, IVW) chosen via Rucker’s framework. Odds ratios (ORs) with 95% confidence intervals (CIs) displayed.

**S3 Figure. (A) Non-linear Mendelian randomisation (MR) estimates for relationships between BMI and female reproductive disorders**. Localised average causal estimates (LACE) are calculated by dividing the IV-free exposure into 100 quantiles (fractional polynomial method) or 10 quantiles (piecewise linear method) with the odds ratio (OR) and 95% confidence intervals (CI) displayed. The reference point for OR = 1 is mean BMI, 27.0 kg/m^2^. **(B) Heterogeneity across BMI spectrum in instrument variables used for non-linear MR**. Instrument variable (IV)-free BMI was divided into quantiles as described in (A). Left: fractional polynomial 100 quantiles, right: piecewise linear 10 quantiles. Proportion of variance in BMI explained by instrument SNPs in each quantile is plotted (β), with 95% confidence intervals shown as standard error bars. Spont. misc. = spontaneous miscarriage.

**S4 Figure. Single-SNP genetic effect estimates for obesity instruments on female reproductive disorders**. SNPs are annotated with their nearest gene by SNPsnap and clustered by obesity-female reproductive disorder relationship. SNPs with >= 80% probability of belonging to a substantial cluster (MRClust) are displayed, with an * if the SNP belongs to substantial clusters for multiple disorders. Effect size estimates are scaled to a variance of 1 within each disease.

**S1 Table. Supplemental Tables A-P**.

## References

1. Wei S, Schmidt MD, Dwyer T, Norman RJ, Venn AJ. Obesity and menstrual irregularity: associations with SHBG, testosterone, and insulin. Obesity (Silver Spring). 2009;17(5):1070–6.

2. Douchi T, Kuwahata R, Yamamoto S, Oki T, Yamasaki H, Nagata Y. Relationship of upper body obesity to menstrual disorders. Acta Obstet Gynecol Scand. 2002;81(2):147–50.

3. Gallagher CS, Makinen N, Harris HR, Rahmioglu N, Uimari O, Cook JP, et al. Genome-wide association and epidemiological analyses reveal common genetic origins between uterine leiomyomata and endometriosis. Nat Commun. 2019;10(1):4857.

4. Yi KW, Shin JH, Park MS, Kim T, Kim SH, Hur JY. Association of body mass index with severity of endometriosis in Korean women. Int J Gynaecol Obstet. 2009;105(1):39–42.

5. Diamanti-Kandarakis E. Role of obesity and adiposity in polycystic ovary syndrome. Int J Obes (Lond). 2007;31 Suppl 2:S8-13; discussion S31-2.

6. Glueck CJ, Goldenberg N. Characteristics of obesity in polycystic ovary syndrome: Etiology, treatment, and genetics. Metabolism. 2019;92:108–20.

7. Spradley FT. Metabolic abnormalities and obesity’s impact on the risk for developing preeclampsia. Am J Physiol Regul Integr Comp Physiol. 2017;312(1):R5–R12.

8. Lashen H, Fear K, Sturdee DW. Obesity is associated with increased risk of first trimester and recurrent miscarriage: matched case-control study. Hum Reprod. 2004;19(7):1644–6.

9. Metwally M, Saravelos SH, Ledger WL, Li TC. Body mass index and risk of miscarriage in women with recurrent miscarriage. Fertil Steril. 2010;94(1):290–5.

10. van der Steeg JW, Steures P, Eijkemans MJ, Habbema JD, Hompes PG, Burggraaff JM, et al. Obesity affects spontaneous pregnancy chances in subfertile, ovulatory women. Hum Reprod. 2008;23(2):324–8.

11. Wise LA, Rothman KJ, Mikkelsen EM, Sorensen HT, Riis A, Hatch EE. An internet-based prospective study of body size and time-to-pregnancy. Hum Reprod. 2010;25(1):253–64.

12. Grodstein F, Goldman MB, Cramer DW. Body mass index and ovulatory infertility. Epidemiology. 1994;5(2):247–50.

13. Missmer SA, Hankinson SE, Spiegelman D, Barbieri RL, Marshall LM, Hunter DJ. Incidence of laparoscopically confirmed endometriosis by demographic, anthropometric, and lifestyle factors. Am J Epidemiol. 2004;160(8):784–96.

14. Shah DK, Correia KF, Vitonis AF, Missmer SA. Body size and endometriosis: results from 20 years of follow-up within the Nurses’ Health Study II prospective cohort. Hum Reprod. 2013;28(7):1783–92.

15. Dixon SC, Nagle CM, Thrift AP, Pharoah PD, Pearce CL, Zheng W, et al. Adult body mass index and risk of ovarian cancer by subtype: a Mendelian randomization study. Int J Epidemiol. 2016;45(3):884–95.

16. Painter JN, O’Mara TA, Marquart L, Webb PM, Attia J, Medland SE, et al. Genetic Risk Score Mendelian Randomization Shows that Obesity Measured as Body Mass Index, but not Waist:Hip Ratio, Is Causal for Endometrial Cancer. Cancer Epidemiol Biomarkers Prev. 2016;25(11):1503–10.

17. Brower MA, Hai Y, Jones MR, Guo X, Chen YI, Rotter JI, et al. Bidirectional Mendelian randomization to explore the causal relationships between body mass index and polycystic ovary syndrome. Hum Reprod. 2019;34(1):127–36.

18. Mahutte NG, Matalliotakis IM, Goumenou AG, Vassiliadis S, Koumantakis GE, Arici A. Inverse correlation between peritoneal fluid leptin concentrations and the extent of endometriosis. Hum Reprod. 2003;18(6):1205–9.

19. Markowska A, Rucinski M, Drews K, Malendowicz LK. Further studies on leptin and leptin receptor expression in myometrium and uterine myomas. Eur J Gynaecol Oncol. 2005;26(5):517–25.

20. Brannian JD, Schmidt SM, Kreger DO, Hansen KA. Baseline non-fasting serum leptin concentration to body mass index ratio is predictive of IVF outcomes. Hum Reprod. 2001;16(9):1819–26.

21. Plowden TC, Zarek SM, Rafique S, Sjaarda LA, Schisterman EF, Silver RM, et al. Preconception leptin levels and pregnancy outcomes: A prospective cohort study. Obes Sci Pract. 2020;6(2):181–8.

22. AlAshqar A, Patzkowsky K, Afrin S, Wild R, Taylor HS, Borahay MA. Cardiometabolic Risk Factors and Benign Gynecologic Disorders. Obstet Gynecol Surv. 2019;74(11):661–73.

23. Hauth JC, Clifton RG, Roberts JM, Myatt L, Spong CY, Leveno KJ, et al. Maternal insulin resistance and preeclampsia. Am J Obstet Gynecol. 2011;204(4):327 e1–6.

24. Butler MG, McGuire A, Manzardo AM. Clinically relevant known and candidate genes for obesity and their overlap with human infertility and reproduction. J Assist Reprod Genet. 2015;32(4):495–508.

25. Sudlow C, Gallacher J, Allen N, Beral V, Burton P, Danesh J, et al. UK biobank: an open access resource for identifying the causes of a wide range of complex diseases of middle and old age. PLoS Med. 2015;12(3):e1001779.

26. Glickman ME, Rao SR, Schultz MR. False discovery rate control is a recommended alternative to Bonferroni-type adjustments in health studies. J Clin Epidemiol. 2014;67(8):850–7.

27. Carreras-Torres R, Johansson M, Haycock PC, Relton CL, Davey Smith G, Brennan P, et al. Role of obesity in smoking behaviour: Mendelian randomisation study in UK Biobank. BMJ. 2018;361:k1767.

28. Ambler GB, Axel. mfp: Multivariable Fractional Polynomials. In: Luecke S, editor. 1.5.2 ed: CRAN; 2015.

29. Wood S. mgcv: Mixed GAM Computation Vehicle with Automatic Smoothness Estimation. In: Wood S, editor. 1.8-31 ed: CRAN; 2021.

30. H. A. Information Theory and an Extension of the Maximum Likelihood Principle. In: Parzen E. Tk, Kitagawa G., editor. Selected Papers of Hirotugu Akaike. Springer Series in Statistics (Perspectives in Statistics). New York, NY: Springer; 1998.

31. Pulit SL, Stoneman C, Morris AP, Wood AR, Glastonbury CA, Tyrrell J, et al. Meta-analysis of genome-wide association studies for body fat distribution in 694 649 individuals of European ancestry. Hum Mol Genet. 2019;28(1):166–74.

32. Karlsson T, Rask-Andersen M, Pan G, Hoglund J, Wadelius C, Ek WE, et al. Contribution of genetics to visceral adiposity and its relation to cardiovascular and metabolic disease. Nat Med. 2019;25(9):1390–5.

33. Elsworth B, Lyon M, Alexander T, Liu Y, Matthews P, Hallett J, et al. The MRC IEU OpenGWAS data infrastructure. bioRxiv. 2020:2020.08.10.244293.

34. Lotta LA, Wittemans LBL, Zuber V, Stewart ID, Sharp SJ, Luan J, et al. Association of Genetic Variants Related to Gluteofemoral vs Abdominal Fat Distribution With Type 2 Diabetes, Coronary Disease, and Cardiovascular Risk Factors. JAMA. 2018;320(24):2553–63.

35. Munafo MR, Tilling K, Taylor AE, Evans DM, Davey Smith G. Collider scope: when selection bias can substantially influence observed associations. Int J Epidemiol. 2018;47(1):226–35.

36. Pirastu N, Cordioli M, Nandakumar P, Mignogna G, Abdellaoui A, Hollis B, et al. Genetic analyses identify widespread sex-differential participation bias. bioRxiv. 2021:2020.03.22.001453.

37. Zhou W, Nielsen JB, Fritsche LG, Dey R, Gabrielsen ME, Wolford BN, et al. Efficiently controlling for case-control imbalance and sample relatedness in large-scale genetic association studies. Nat Genet. 2018;50(9):1335–41.

38. Willer CJ, Li Y, Abecasis GR. METAL: fast and efficient meta-analysis of genomewide association scans. Bioinformatics. 2010;26(17):2190–1.

39. Painter JN, Anderson CA, Nyholt DR, Macgregor S, Lin J, Lee SH, et al. Genome-wide association study identifies a locus at 7p15.2 associated with endometriosis. Nat Genet. 2011;43(1):51–4.

40. Laisk T, Soares ALG, Ferreira T, Painter JN, Censin JC, Laber S, et al. The genetic architecture of sporadic and multiple consecutive miscarriage. Nat Commun. 2020;11(1):5980.

41. Deng L, Zhang H, Yu K. Power calculation for the general two-sample Mendelian randomization analysis. Genet Epidemiol. 2020;44(3):290–9.

42. Brion MJ, Shakhbazov K, Visscher PM. Calculating statistical power in Mendelian randomization studies. Int J Epidemiol. 2013;42(5):1497–501.

43. Hemani G, Zheng J, Elsworth B, Wade KH, Haberland V, Baird D, et al. The MR-Base platform supports systematic causal inference across the human phenome. Elife. 2018;7.

44. Bowden J, Del Greco MF, Minelli C, Davey Smith G, Sheehan N, Thompson J. A framework for the investigation of pleiotropy in two-sample summary data Mendelian randomization. Stat Med. 2017;36(11):1783–802.

45. Bowden J, Spiller W, Del Greco MF, Sheehan N, Thompson J, Minelli C, et al. Improving the visualization, interpretation and analysis of two-sample summary data Mendelian randomization via the Radial plot and Radial regression. Int J Epidemiol. 2018;47(4):1264–78.

46. Sapkota Y, Steinthorsdottir V, Morris AP, Fassbender A, Rahmioglu N, De Vivo I, et al. Meta-analysis identifies five novel loci associated with endometriosis highlighting key genes involved in hormone metabolism. Nat Commun. 2017;8:15539.

47. Day F, Karaderi T, Jones MR, Meun C, He C, Drong A, et al. Large-scale genome-wide meta-analysis of polycystic ovary syndrome suggests shared genetic architecture for different diagnosis criteria. PLoS Genet. 2018;14(12):e1007813.

48. Bycroft C, Freeman C, Petkova D, Band G, Elliott LT, Sharp K, et al. The UK Biobank resource with deep phenotyping and genomic data. Nature. 2018;562(7726):203–9.

49. Staley JR, Burgess S. Semiparametric methods for estimation of a nonlinear exposure-outcome relationship using instrumental variables with application to Mendelian randomization. Genet Epidemiol. 2017;41(4):341–52.

50. Carter AR, Sanderson E, Hammerton G, Richmond RC, Smith GD, Heron J, et al. Mendelian randomisation for mediation analysis: current methods and challenges for implementation. bioRxiv. 2020:835819.

51. Kilpelainen TO, Carli JF, Skowronski AA, Sun Q, Kriebel J, Feitosa MF, et al. Genome-wide meta-analysis uncovers novel loci influencing circulating leptin levels. Nat Commun. 2016;7:10494.

52. Manning AK, Hivert MF, Scott RA, Grimsby JL, Bouatia-Naji N, Chen H, et al. A genome-wide approach accounting for body mass index identifies genetic variants influencing fasting glycemic traits and insulin resistance. Nat Genet. 2012;44(6):659–69.

53. Walford GA, Gustafsson S, Rybin D, Stancakova A, Chen H, Liu CT, et al. Genome-Wide Association Study of the Modified Stumvoll Insulin Sensitivity Index Identifies BCL2 and FAM19A2 as Novel Insulin Sensitivity Loci. Diabetes. 2016;65(10):3200–11.

54. Hemani G, Tilling K, Davey Smith G. Orienting the causal relationship between imprecisely measured traits using GWAS summary data. PLoS Genet. 2017;13(11):e1007081.

55. Doob JL. The Limiting Distributions of Certain Statistics. The Annals of Mathematical Statistics. 1935;6(3):160-9, 10.

56. Pers TH, Timshel P, Hirschhorn JN. SNPsnap: a Web-based tool for identification and annotation of matched SNPs. Bioinformatics. 2015;31(3):418–20.

57. Foley CN, Mason AM, Kirk PDW, Burgess S. MR-Clust: clustering of genetic variants in Mendelian randomization with similar causal estimates. Bioinformatics. 2021;37(4):531–41.

58. Burgess S, Thompson DJ, Rees JMB, Day FR, Perry JR, Ong KK. Dissecting Causal Pathways Using Mendelian Randomization with Summarized Genetic Data: Application to Age at Menarche and Risk of Breast Cancer. Genetics. 2017;207(2):481–7.

59. Relton CL, Davey Smith G. Two-step epigenetic Mendelian randomization: a strategy for establishing the causal role of epigenetic processes in pathways to disease. Int J Epidemiol. 2012;41(1):161–76.

60. Perry JR, Corre T, Esko T, Chasman DI, Fischer K, Franceschini N, et al. A genome-wide association study of early menopause and the combined impact of identified variants. Hum Mol Genet. 2013;22(7):1465–72.

61. Jones MR, Mathur R, Cui J, Guo X, Azziz R, Goodarzi MO. Independent confirmation of association between metabolic phenotypes of polycystic ovary syndrome and variation in the type 6 17beta-hydroxysteroid dehydrogenase gene. J Clin Endocrinol Metab. 2009;94(12):5034–8.

62. Ortega-Azorin C, Coltell O, Asensio EM, Sorli JV, Gonzalez JI, Portoles O, et al. Candidate Gene and Genome-Wide Association Studies for Circulating Leptin Levels Reveal Population and Sex-Specific Associations in High Cardiovascular Risk Mediterranean Subjects. Nutrients. 2019;11(11).

63. Miranda-Lora AL, Cruz M, Aguirre-Hernandez J, Molina-Diaz M, Gutierrez J, Flores-Huerta S, et al. Exploring single nucleotide polymorphisms previously related to obesity and metabolic traits in pediatric-onset type 2 diabetes. Acta Diabetol. 2017;54(7):653–62.

64. Nielsen TR, Appel EV, Svendstrup M, Ohrt JD, Dahl M, Fonvig CE, et al. A genome-wide association study of thyroid stimulating hormone and free thyroxine in Danish children and adolescents. PLoS One. 2017;12(3):e0174204.

65. Ramly B, Afiqah-Aleng N, Mohamed-Hussein ZA. Protein-Protein Interaction Network Analysis Reveals Several Diseases Highly Associated with Polycystic Ovarian Syndrome. Int J Mol Sci. 2019;20(12).

66. Wu TI, Huang RL, Su PH, Mao SP, Wu CH, Lai HC. Ovarian cancer detection by DNA methylation in cervical scrapings. Clin Epigenetics. 2019;11(1):166.

67. Makabe T, Arai E, Hirano T, Ito N, Fukamachi Y, Takahashi Y, et al. Genome-wide DNA methylation profile of early-onset endometrial cancer: its correlation with genetic aberrations and comparison with late-onset endometrial cancer. Carcinogenesis. 2019;40(5):611–23.

68. Barber TM, Franks S. Obesity and polycystic ovary syndrome. Clin Endocrinol (Oxf). 2021.

69. Teede HJ, Joham AE, Paul E, Moran LJ, Loxton D, Jolley D, et al. Longitudinal weight gain in women identified with polycystic ovary syndrome: results of an observational study in young women. Obesity (Silver Spring). 2013;21(8):1526–32.

70. Norgaard M, Ehrenstein V, Vandenbroucke JP. Confounding in observational studies based on large health care databases: problems and potential solutions - a primer for the clinician. Clin Epidemiol. 2017;9:185–93.

71. VanderWeele TJ, Hernan MA, Robins JM. Causal directed acyclic graphs and the direction of unmeasured confounding bias. Epidemiology. 2008;19(5):720–8.

72. Ezeh U, Yildiz BO, Azziz R. Referral bias in defining the phenotype and prevalence of obesity in polycystic ovary syndrome. J Clin Endocrinol Metab. 2013;98(6):E1088–96.

73. Luque-Ramirez M, Alpanes M, Sanchon R, Fernandez-Duran E, Ortiz-Flores AE, Escobar-Morreale HF. Referral bias in female functional hyperandrogenism and polycystic ovary syndrome. Eur J Endocrinol. 2015;173(5):603–10.

74. Karvonen-Gutierrez C, Kim C. Association of Mid-Life Changes in Body Size, Body Composition and Obesity Status with the Menopausal Transition. Healthcare (Basel). 2016;4(3).

75. McCann SE, Freudenheim JL, Darrow SL, Batt RE, Zielezny MA. Endometriosis and body fat distribution. Obstet Gynecol. 1993;82(4 Pt 1):545–9.

76. Sun K, Xie Y, Zhao N, Li Z. A case-control study of the relationship between visceral fat and development of uterine fibroids. Exp Ther Med. 2019;18(1):404–10.

77. Sato F, Nishi M, Kudo R, Miyake H. Body fat distribution and uterine leiomyomas. J Epidemiol. 1998;8(3):176–80.

78. Lord J, Thomas R, Fox B, Acharya U, Wilkin T. The central issue? Visceral fat mass is a good marker of insulin resistance and metabolic disturbance in women with polycystic ovary syndrome. BJOG. 2006;113(10):1203–9.

79. Ferrero S, Anserini P, Remorgida V, Ragni N. Body mass index in endometriosis. Eur J Obstet Gynecol Reprod Biol. 2005;121(1):94–8.

80. Hediger ML, Hartnett HJ, Louis GM. Association of endometriosis with body size and figure. Fertil Steril. 2005;84(5):1366–74.

81. Berlanda N, Somigliana E, Frattaruolo MP, Buggio L, Dridi D, Vercellini P. Surgery versus hormonal therapy for deep endometriosis: is it a choice of the physician? Eur J Obstet Gynecol Reprod Biol. 2017;209:67–71.

82. Kim SA, Um MJ, Kim HK, Kim SJ, Moon SJ, Jung H. Study of dienogest for dysmenorrhea and pelvic pain associated with endometriosis. Obstet Gynecol Sci. 2016;59(6):506–11.

83. Jeng CJ, Chuang L, Shen J. A comparison of progestogens or oral contraceptives and gonadotropin-releasing hormone agonists for the treatment of endometriosis: a systematic review. Expert Opin Pharmacother. 2014;15(6):767–73.

84. Okifuji A, Hare BD. The association between chronic pain and obesity. J Pain Res. 2015;8:399–408.

85. Engstrom G, Hedblad B, Stavenow L, Lind P, Janzon L, Lindgarde F. Inflammation-sensitive plasma proteins are associated with future weight gain. Diabetes. 2003;52(8):2097–101.

86. Yasui T, Hayashi K, Mizunuma H, Kubota T, Aso T, Matsumura Y, et al. Association of endometriosis-related infertility with age at menopause. Maturitas. 2011;69(3):279–83.

87. Sirmans SM, Pate KA. Epidemiology, diagnosis, and management of polycystic ovary syndrome. Clin Epidemiol. 2013;6:1–13.

88. Laven JS, Imani B, Eijkemans MJ, Fauser BC. New approach to polycystic ovary syndrome and other forms of anovulatory infertility. Obstet Gynecol Surv. 2002;57(11):755–67.

89. Mardh PA. Tubal factor infertility, with special regard to chlamydial salpingitis. Curr Opin Infect Dis. 2004;17(1):49–52.

90. de Ziegler D, Borghese B, Chapron C. Endometriosis and infertility: pathophysiology and management. Lancet. 2010;376(9742):730–8.

91. Homer HA. The Role of Oocyte Quality in Explaining “Unexplained” Infertility. Semin Reprod Med. 2020;38(1):21–8.

92. Arojoki M, Jokimaa V, Juuti A, Koskinen P, Irjala K, Anttila L. Hypothyroidism among infertile women in Finland. Gynecol Endocrinol. 2000;14(2):127–31.

93. Guerin LR, Prins JR, Robertson SA. Regulatory T-cells and immune tolerance in pregnancy: a new target for infertility treatment? Hum Reprod Update. 2009;15(5):517–35.

94. Luciano AA, Lanzone A, Goverde AJ. Management of female infertility from hormonal causes. Int J Gynaecol Obstet. 2013;123 Suppl 2:S9–17.

95. Sen A, Kushnir VA, Barad DH, Gleicher N. Endocrine autoimmune diseases and female infertility. Nat Rev Endocrinol. 2014;10(1):37–50.

96. Ramlau-Hansen CH, Thulstrup AM, Nohr EA, Bonde JP, Sorensen TI, Olsen J. Subfecundity in overweight and obese couples. Hum Reprod. 2007;22(6):1634–7.

97. Nilufer R, Karina B, Paraskevi C, Rebecca D, Genevieve G, Ayush G, et al. Large-scale genome-wide association meta-analysis of endometriosis reveals 13 novel loci and genetically-associated comorbidity with other pain conditions. bioRxiv. 2018:406967.

98. Catherino WH, Eltoukhi HM, Al-Hendy A. Racial and ethnic differences in the pathogenesis and clinical manifestations of uterine leiomyoma. Semin Reprod Med. 2013;31(5):370–9.

99. Marsh EE, Ekpo GE, Cardozo ER, Brocks M, Dune T, Cohen LS. Racial differences in fibroid prevalence and ultrasound findings in asymptomatic young women (18-30 years old): a pilot study. Fertil Steril. 2013;99(7):1951–7.

100. Bougie O, Healey J, Singh SS. Behind the times: revisiting endometriosis and race. Am J Obstet Gynecol. 2019;221(1):35 e1–e5.

101. Kim JJ, Choi YM. Phenotype and genotype of polycystic ovary syndrome in Asia: Ethnic differences. J Obstet Gynaecol Res. 2019;45(12):2330–7.

102. Nakimuli A, Chazara O, Byamugisha J, Elliott AM, Kaleebu P, Mirembe F, et al. Pregnancy, parturition and preeclampsia in women of African ancestry. Am J Obstet Gynecol. 2014;210(6):510–20 e1.

103. Tan X, Li S, Chang Y, Fang C, Liu H, Zhang X, et al. Effect of metformin treatment during pregnancy on women with PCOS: a systematic review and meta-analysis. Clin Invest Med. 2016;39(4):E120–31.

