## Supplemental Figure 1 for "The role of obesity in female reproductive conditions: A Mendelian Randomisation study"

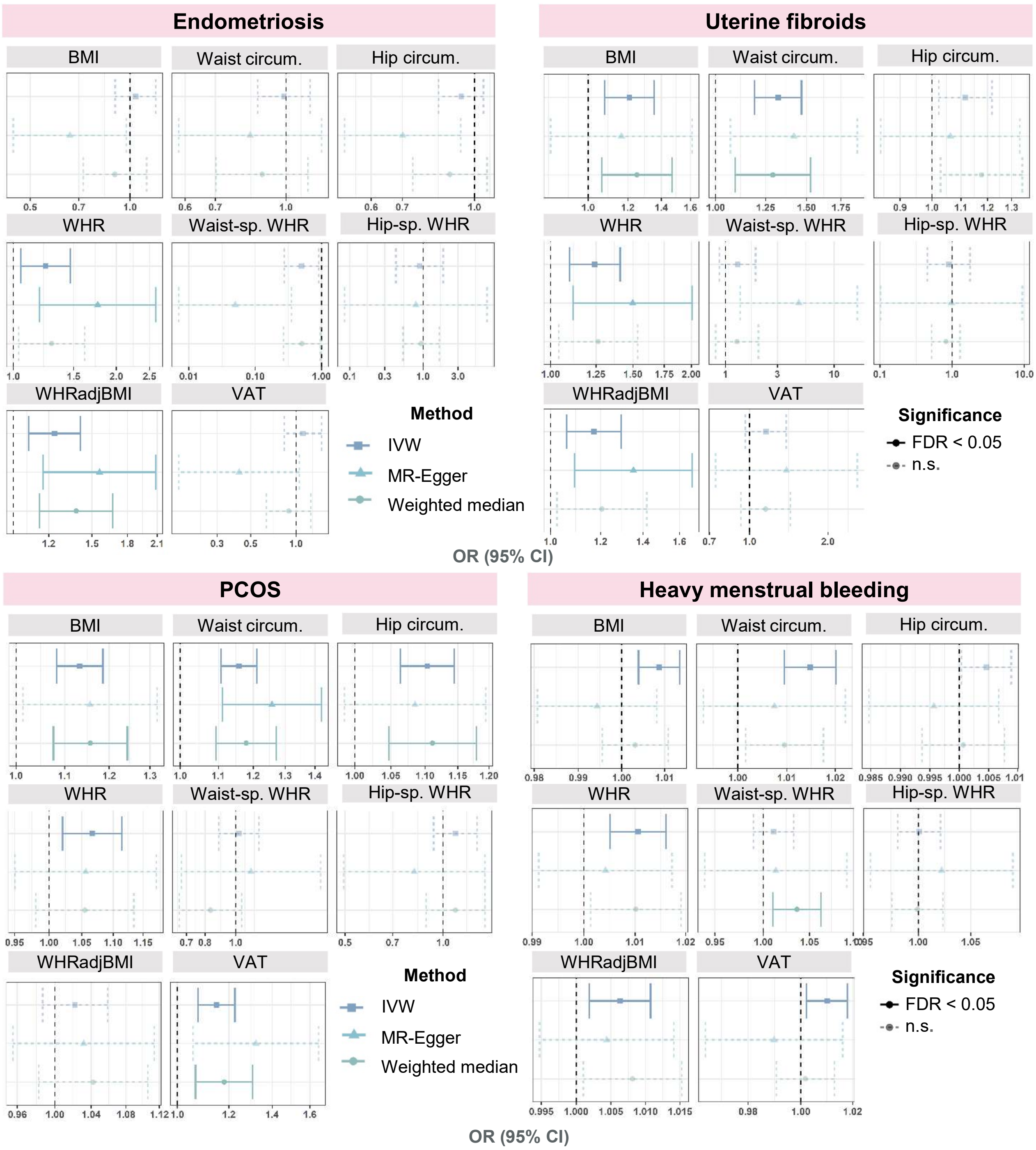


**S1 Figure. Comparison of different Mendelian randomisation methods for regression of female reproductive diseases on various obesity traits.** Summary statistics MR was performed with the R package TwoSampleMR using three different methods whose results are compared. Effect sizes are displayed as odds ratios (ORs) with 95% confidence intervals (CIs). P-values are adjusted for multiple testing with the false discovery rate (FDR) correction; solid lines indicate associations that are significant at an adjusted p-value threshold of 0.05. BMI = body mass index, IVW = Inverse-variance weighted, PCOS = polycystic ovary syndrome, WHR = waist-hip ratio, WHRadjBMI = WHR adjusted for BMI, VAT = genetically predicted visceral adipose tissue mass… contd.

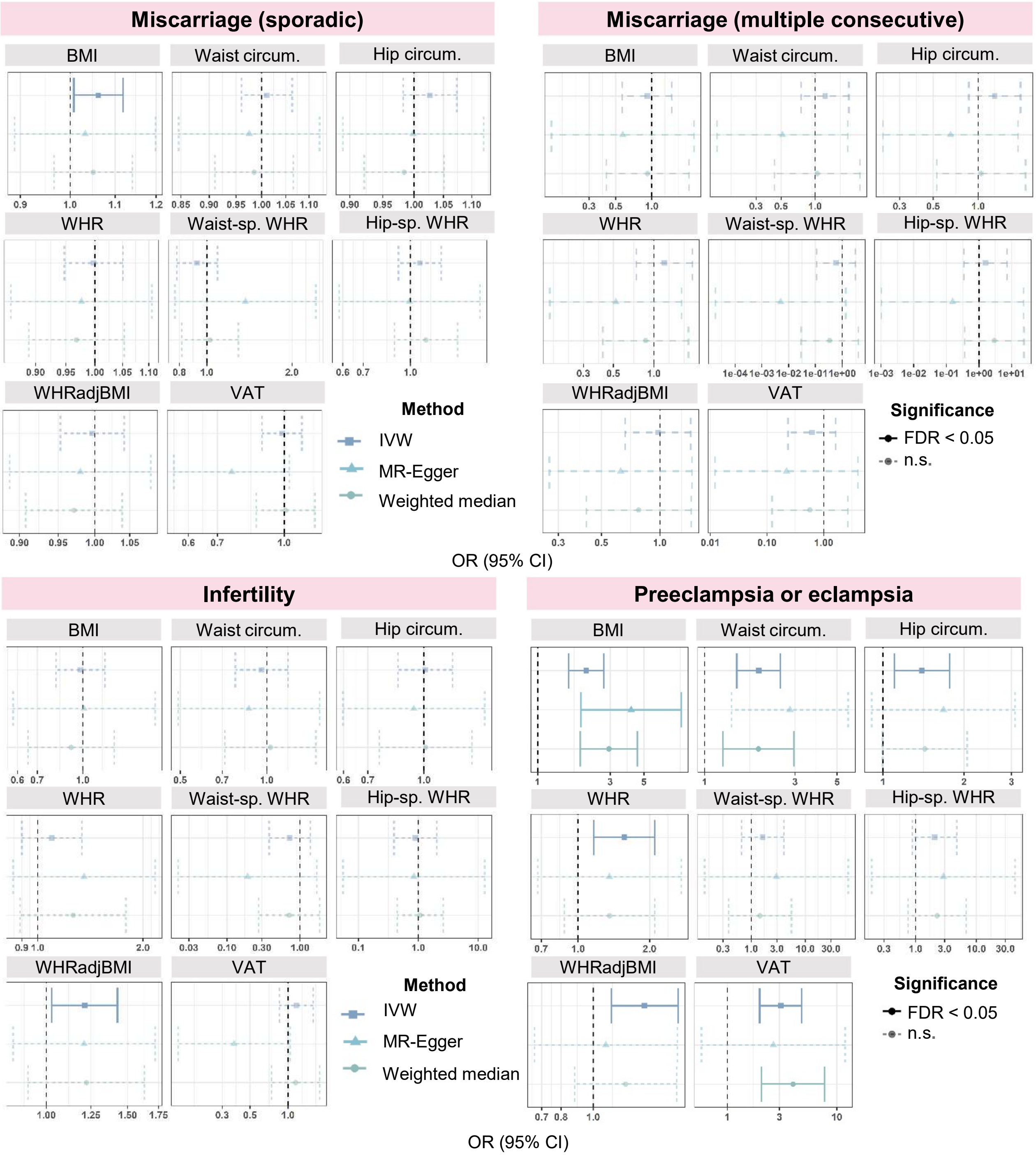


**… contd. S1 Figure. Comparison of different Mendelian randomisation methods for regression of female reproductive diseases on various obesity traits.**
