## Supplemental Figure 2 for "The role of obesity in female reproductive conditions: A Mendelian Randomisation study"

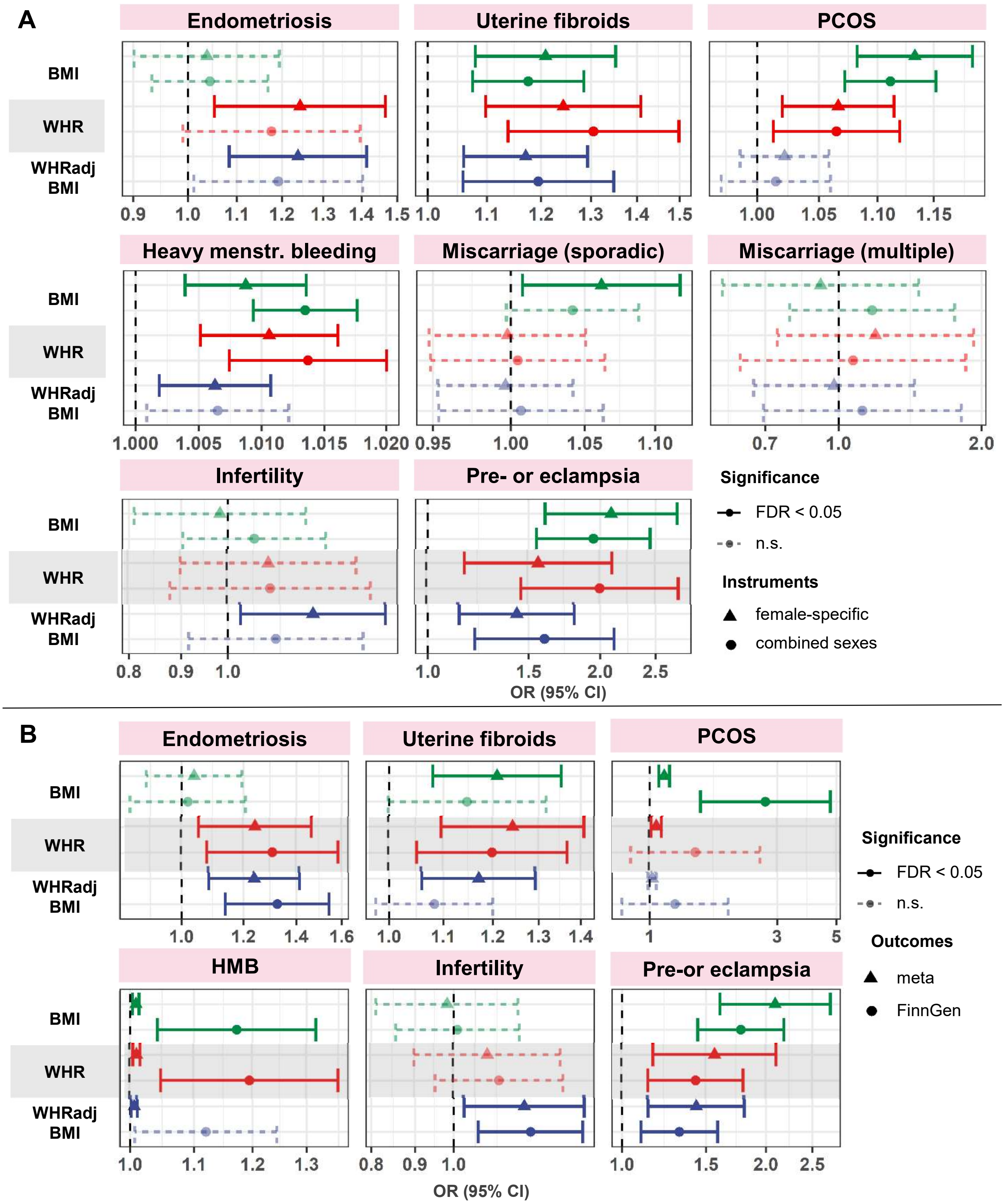


**S2 Figure. Mendelian randomisation sensitivity analyses for female reproductive disorders regressed on obesity traits. (A)** Female-specific genetic instruments vs combined-sexes genetic instruments. **(B)** Reproductive outcomes from meta-analysis of UKBB and FinnGen summary statistics vs those from FinnGen only. Summary statistics MR performed with TwoSampleMR R package and best method (displayed, IVW) chosen via Rucker's framework. Odds ratios (ORs) with 95% confidence intervals (CIs) displayed.
