## Supplemental Figure 3 for "The role of obesity in female reproductive conditions: A Mendelian Randomisation study"

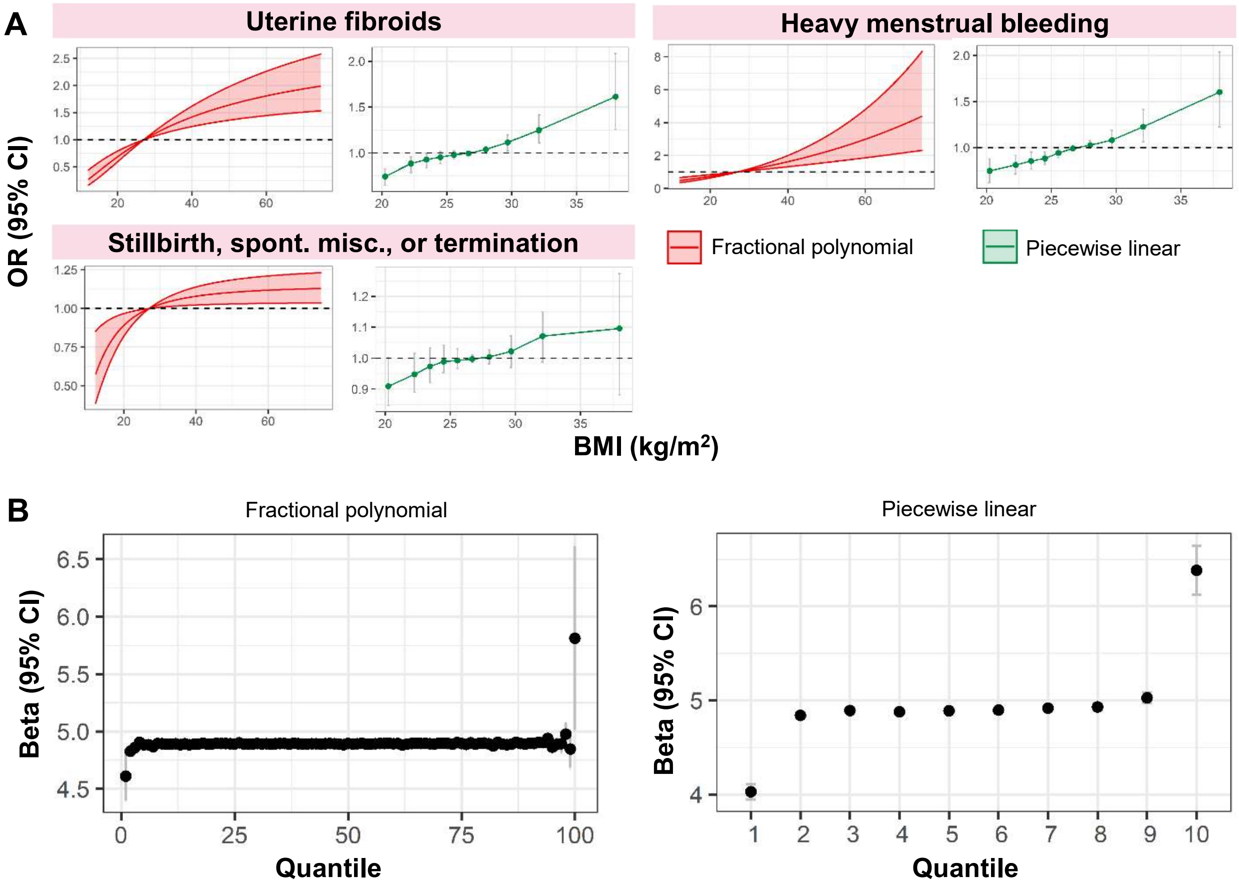


**S3 Figure. (A) Non-linear Mendelian randomisation (MR) estimates for relationships between BMI and female reproductive disorders.** Localised average causal estimates (LACE) are calculated by dividing the IV-free exposure into 100 quantiles (fractional polynomial method) or 10 quantiles (piecewise linear method) with the odds ratio (OR) and 95% confidence intervals (CI) displayed. The reference point for OR = 1 is mean BMI, 27.0 kg/$m^2$. **(B) Heterogeneity across BMI spectrum in instrument variables used for non-linear MR.** Instrument variable (IV)-free BMI was divided into quantiles as described in (A). Left: fractional polynomial 100 quantiles, right: piecewise linear 10 quantiles. Proportion of variance in BMI explained by instrument SNPs in each quantile is plotted (β), with 95% confidence intervals shown as standard error bars.
Spont. misc. = spontaneous miscarriage.
