## Supplemental Figure 4 for "The role of obesity in female reproductive conditions: A Mendelian Randomisation study"

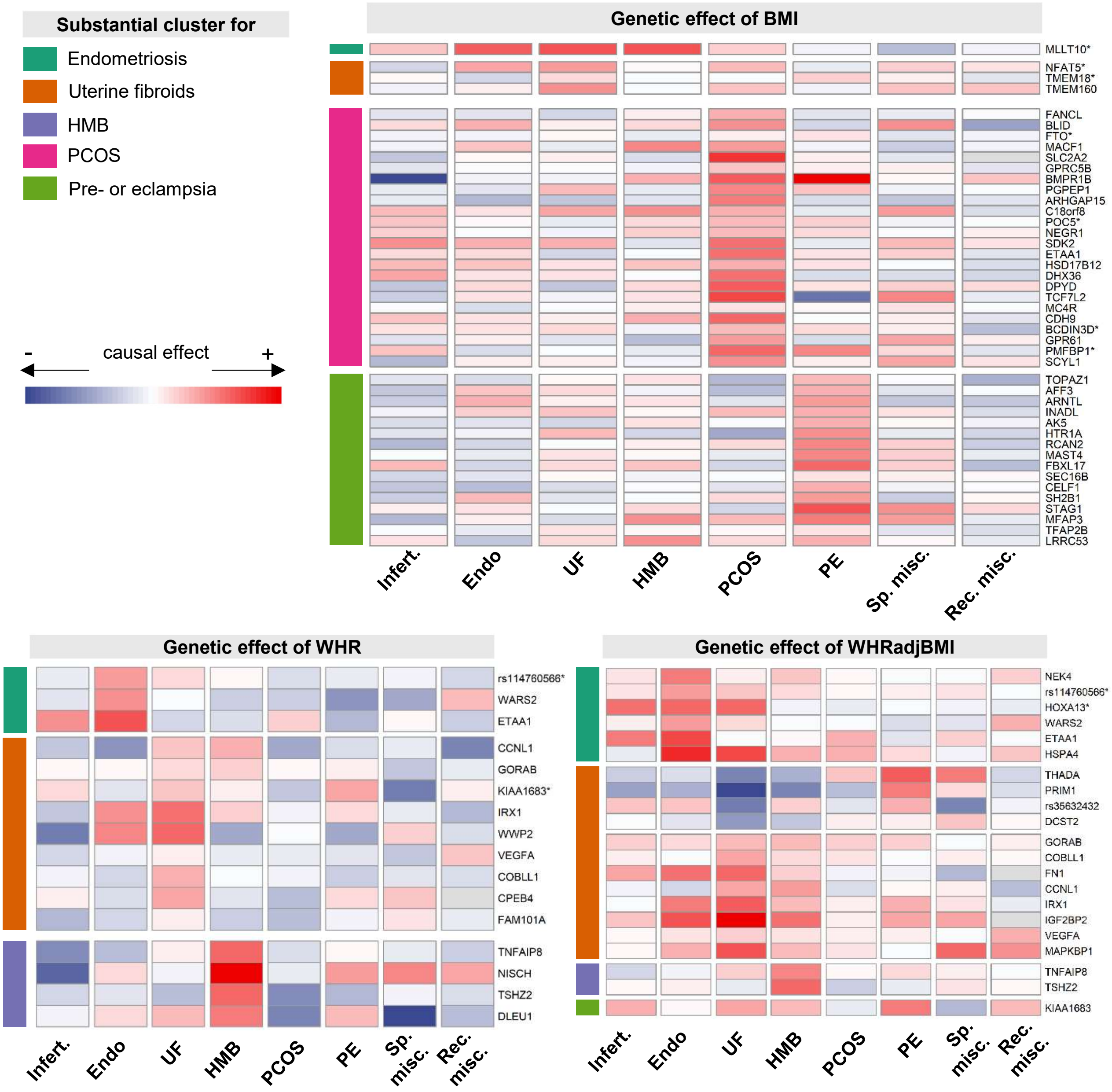


**S4 Figure. Single-SNP genetic effect estimates for obesity instruments on female reproductive disorders.** SNPs are annotated with their nearest gene by SNPsnap and clustered by obesity-female reproductive disorder relationship. SNPs with >= 80% probability of belonging to a substantial cluster (MRClust) are displayed, with an * if the SNP belongs to substantial clusters for multiple disorders. Effect size estimates are scaled to a variance of 1 within each disease.
